# Automated abstraction of clinical parameters of multiple myeloma from real-world clinical notes using large language models

**DOI:** 10.1101/2024.12.17.24318605

**Authors:** Alana O’Brien Del Campo, Dmytro Lituiev, Gowtham Varma, Mithun Manoharan, Sunil Kumar Ravi, Avinash Aman, Ankit Kansagra, Joel Greshock, AJ Venkatakrishnan, Ashita Batavia

## Abstract

**Background:** Real-world evidence (RWE) is increasingly recognized as a valuable type of oncology research but extracting fit-for-purpose real-world data (RWD) from electronic health records (EHRs) remains challenging. Manual abstraction from free-text clinical documents, although the gold standard for information extraction, is resource-intensive. RWD generation using natural language processing (NLP) has been limited by performance ceilings and annotation requirements, which recent LLMs improve on. We evaluate new NLP workflows in abstracting multiple myeloma (MM) information from de-identified EHRs.

**Methods:** NLP workflows (BERT and Llama-based using various prompt types) were developed for 12 MM-specific data fields and evaluated with manually curated data from 125 clinical notes. The best Llama-based workflow for three data fields was applied to all recent notes in selected charts to generate patient journey timelines.

**Results:** Average F_1_ for the best Llama and BERT workflows was 0.82 and 0.65 respectively. Best workflow performance ranged across the data fields (F_1_ = 0.59–0.99). Statistical analysis of the results showed model size, inter-rater reliability (IRR), variable type, and prompt design significantly predicted workflow performance, in descending order of significance (*p* < 0.05).

**Conclusion:** The overall performance improvements seen with larger LLMs and chain-of-thought prompting was greater in ambiguous data fields. IRR can be used to prioritize NLP resources and increase efficiency of RWD generation without sacrificing data quality.

## Introduction

In the past decade, multiple myeloma (MM) patients and providers have seen a rapid expansion of therapeutic options, creating greater need for real world evidence (RWE) that can expand on findings from randomized controlled trials.^1–4^ Electronic Health Records (EHRs) contain a wealth of real-world clinical data capturing diagnoses, treatments and outcomes. However, key concepts for creating trial-like cohorts and outcomes, such as transplant eligibility and certain International Myeloma Working Group (IMWG) criteria, are captured in unstructured clinical notes and must be extracted and structured to create fit-for-purpose real world data (RWD) usable for evidence generation.^5,6^

Manual abstraction continues to be the gold standard for clinical information extraction (IE) from unstructured text. However, the time and resources required to manually abstract numerous clinical data fields may ultimately constrain RWE generation. In recent years, natural language processing (NLP) techniques based on transformer language models have shown great potential for automating this labor-intensive process.^7^ Transformer models can be broadly categorized into encoders such as Bidirectional Encoder Representations from Transformer (BERT) typically used for discriminative tasks like classification and named entity recognition (NER), and decoders (such as GPT and Llama) and encoder-decoders typically used for tasks requiring text generation. Recent decoder models typically have at least 1 billion parameters compared to earlier decoder models (e.g., BERT with 300 million parameters).

Until recently, NER-based IE using BERT models has been a popular NLP approach due to their strong performance across many tasks.^8–10^ The effectiveness of BERT models for clinical IE has previously been demonstrated in tasks like identifying drug-related adverse events, extracting clinical symptoms and real-world disease outcomes.^11–13^ Their success notwithstanding, BERT models pose certain limitations to investigators. BERT models, while highly parameterized compared to traditional NLP techniques, still have fewer parameters than most of the newer LLMs and often require task-specific training on manually labeled datasets.^14,15^ Also, the narrower context window of most BERT models limits their ability to infer concepts that are typically presented over multiple sentences or paragraphs, such as transplant eligibility.^16^

More recent generative LLMs have more parameters and larger context windows. When pre-trained on large corpora, these newer LLMs promise high off-the-shelf performance in clinical IE tasks without task-specific training, particularly for complex concepts.^17^ These advances in LLMs thus hold promise to accelerate development of high quality real-world datasets by mitigating the need for extensive manual data curation and fine-tuning.

Early applications of generative LLMs for clinical IE predominantly use Flan and GPT family models, and demonstrate wide-ranging performance (accuracy and F1 statistics both ranging from low 40s to high 90s percentage), primarily due to three contributing factors.^18–23^ First, complex data fields, such as adverse social determinants of health, challenge all NLP techniques including LLMs.^24,25^ Model size, which impacts pre-training capacity, drives off-the-shelf performance of LLMs. And finally, reasoning-based prompting techniques like chain-of-thought purport to help a generally pre-trained LLM handle specialized tasks better than zero-shot learning.^26,27^

Investigators planning RWD generation from unstructured text can find newer LLMs to be a powerful tool, but the lack of comparability between published studies hinders generalization of results to inform IE strategy. Off-the-shelf accuracy of LLMs is unpredictable, obfuscating the time and effort required to produce a reliable data field. Although larger LLMs achieve better results, they require more time to run, can be costly, and request substantial up-front investment in computational infrastructure. Also, prompt engineering is an art more than a science, with few guardrails to guide users on improving performance.

Instead of defaulting to a single NLP technique for all data fields, efficient data generation balances accuracy versus time and resource demands.^28^ Our objective in this research is quantify the impact of IE design choices and provide investigators with an approach to tailoring efficient LLM usage. Ultimately, we hope this will accelerate generation of high-quality MM RWD.

In this research, we utilize several NLP models (Llama 3 8B, Llama 3 70B, and BERT) in workflows to extract clinical information from the EHR notes of patients diagnosed with MM. We evaluate NLP workflow performance against a manually abstracted reference dataset. We analyze the impact of text ambiguity, model size and chain-of-thought prompting on newer LLM performance. Finally, we use the best NLP workflow to extract three data fields for all recent notes in select patient charts, demonstrating enhanced information availability using the entirety of the observable patient journey.

## Methods

### Data source & selection

This study analyzed de-identified EHR data from a network of tertiary clinical centers tied to an academic medical center in the United States through the nference nSights Analytics Platform.^29^ nference, in collaboration with the academic medical center data partner (AMC) that provided the de-identified data for this study, has established a secure data environment, hosted by and within the AMC, that houses the AMC’s de-identified patient data. The provisioning of and access to this data are governed by an expert determination that satisfies the HIPAA Privacy Rule requirements for the de-identification of protected health information. Each AMC’s de-identified data environment is specifically designed and operated to enable access to and analysis of de-identified data without the need for Institutional Review Board (IRB) oversight, approval, or an exemption confirmation. Given these measures, informed consent and IRB review were not required for this study.

#### Patient population and selected notes

A cohort of MM patients was created and notes for IE were sampled from their charts. The study cohort (n=3,793) included patients with MM diagnosis and treatment between January 1, 2019, to March 31, 2024. Patients with MM were identified using two occurrences of diagnosis codes of 203.0* (ICD-9), C90, and C90.0* (ICD-10). The first occurrence of MM diagnosis code was considered the diagnosis date. Patients were required to have at least one encounter recorded within six months of the diagnosis date and another encounter six months after the diagnosis date.

The notes database (n = 250 notes) was created by selecting one instance of provider-documented unstructured text associated with an encounter, pathology testing, or imaging study, from the charts of 250 patients randomly selected from the study cohort. Study cohort and note selection criteria are available in Supplementary Note 1. This notes database was randomly divided into development and test sets of 125 notes each.

### Clinical concepts and data fields

13 MM-related data fields were chosen. (See **Table 1**). All fields were text-based, provider-documented values (i.e., not derived from structured data fields such as ICD codes, medications, or timestamps).

**Table 1.** Data fields of interest contained in HCP notes.

| **Table 1. Data fields of interest contained in HCP notes** |  |  |
| --- | --- | --- |
| **Concept** | **Data field** | **Value type and range** |
| MM disease state | *1. MM diagnosis* | Categorical<br>*Yes / Likely / No / Unspecified\** |
|  | *2. MM status* | Categorical<br>*Newly diagnosed / Relapsed / Refractory / Remission* |
|  | *3. MM diagnosis date* | Datetime<br>*YYYY-MM-DD* |
| Transplant eligibility & status | *4. Transplant eligibility & status* | Categorical<br>*Eligible / Eligible but deferred / Ineligible / Performed / Unspecified\** |
| Eastern Cooperative Oncology Group (ECOG) performance status scale | *5. ECOG score* | Integer<br>*0—5* |
| Plasmacytosis | *6. Plasmacytosis percentage* | Integer<br>*0-100%* |
| MM-related bone lesions | *7. MM-related bone lesion presence* | Binary<br>*Yes / No / Unrelated / Unspecified\** |
| Extra-medullary disease (EMD) | *8. EMD presence* | Binary<br>*Present / No / Unspecified\** |
|  | *9. EMD location* | Binary<br>*Paramedullary / Soft tissue / Unspecified\** |
| HCP-documented First line therapy (FLT) | *10. FLT regimen* | Categorical<br>*Set of drugs and procedures, mapped to NDC or RxNorm and HCPCS or CPT\*\** |
|  | *11. FLT start date* | Datetime<br>*YYYY-MM-DD* |
| HCP-documented FLT response | *12. FLT response* | Categorical<br>*IMWG 2016 response categories: Stringent Complete Response / Complete Response / Very Good Partial Response / Partial Response / Minimal Response / No Response / Progressive Disease / Unspecified\** |
|  | *13. FLT response date* | Datetime<br>*Formatted as YYYY-MM-DD* |
| *Unspecified indicates no mention in text<br>**NDC: National Drug Code, RxNorm: National Library of Medicine (NLM) issued normalized naming system for generic and branded Drugs. HCPCS: Healthcare Common Procedure Coding System. CPT: Current Procedure Terminology. |  |  |

### Manual data curation

A reference dataset for the 13 data fields was created for prompt development and NLP workflow testing. The reference dataset was created by two independent abstractors with arbitration by a third abstractor.

Inter-rater reliability (IRR) was calculated to approximate the ambiguity of the information available. IRR was assessed in two ways. First, Krippendorff’s α (K-α) was calculated. An overall average K-α of ≥0.8 for the test dataset was considered acceptable.^30^ Secondly, the agreement between each abstractor and the final arbitrated label was evaluated using an F_1_ score, which was then averaged across the two abstractors. This F_1_-based IRR (subsequently referred to as IRR-F_1_) was used as a proxy for data field ambiguity in statistical analysis.

Supplementary Note 2 contains abstraction protocol and metrics (K-α and F_1_ score for each data field, class distribution) and low-count class combinations.

### NLP-based methods for information extraction

Five LLM workflows for extracting information on the selected data fields were developed: four workflows utilizing Llama models and one workflow utilizing BERT.

#### Llama workflows

Meta’s Llama 3 family of models provided an open-source, privately deployable LLM with small and medium-sized model sizes.^31^ The small model (8 billion parameters) has lower performance on all benchmarks assessed at release but is more computationally facile.^31^ The medium model (70 billion parameters) offers advanced natural language capabilities, more promising for challenging clinical text, but required more computational infrastructure to run.^31^ (A 405 billion parameter model was released after the start of research and thus not included.)

The small (8B) and medium (70B) models were run in private compute clusters to protect deidentified clinical text. Performance evaluations were done using a temperature of 0.1 and a top-K (the number of highest probability token options used for sampling) value of 1 for reproducible results. Sensitivity analysis was performance for additional top-K and temperature values.

Zero-shot-learning (ZSL) was selected for default performance and chain-of-thought (CoT) prompting was compared as a common reasoning-method technique. Bespoke prompts for each data field were systematically designed using the development set. Prompts were refined based on errors generated until achieving average abstractor F_1_ for the respective field or when subsequent modifications did not yield performance improvements. Final prompts are available in Supplementary Note 3. The model was prompted to return a JSON-structured response. The syntax of JSON output was parsed and standardized to pre-specified labels using post-processing logic.

These four Llama workflows (i.e., model-prompt combination) are referenced throughout using respective model size and prompting technique (e.g., 70B-CoT for Llama 3 70B with chain-of-thought prompting).

#### BERT workflow

A BERT workflow, earlier developed for a broad range of biomedical tasks on nference data, was deployed as the baseline technique. A pipeline of six proprietary BERT-based classification models includes: (1) named entity recognition model trained to detect 27 entity types; (2-4) qualifier models: subject, temporality and certainty models; (5) concept association model for “problem-location”, “problem-severity”, “lab data-value”; and (6) date association model for “variable-date” entity pairs. These proprietary models are fine-tuned versions of SciBERT cased^32^ (basis for models 1-5 specified above) and ClinicalBERT^33^ (basis for model 6). The base models underwent further supervised fine-tuning for IE tasks on annotated sentences from clinical document texts of the nference nSights database, but not specifically on MM patient note database.

For each data field, relevant synonyms derived from tokenization of the development set were curated and incorporated. The BERT pipeline output was refined using regular expression models and business rules developed using the development set

BERT workflow development approach, performance metrics, and rules are available in Supplementary Note 4.

### Statistical analysis

Macro-F_1_ scores were used to evaluate the performance of the NLP workflows. For multi-label fields such as dates, macro-F_1_ score was substituted with a weighted F_1_, calculated as defined in Supplementary Note 1. Spearman’s rho was calculated for numeric fields. Visualization and exploratory data analysis was conducted in Python (3.10.6) using pandas (2.2.2), numpy (1.26.4), scipy, matplotlib (3.9.1), seaborn (0.13.2), and plotly (5.23.0) (https://plot.ly/).^34–39^

Statistical analysis of LLM workflow performance for each data field included pairwise comparisons and, for the four Llama workflows, ANOVA. Independent variables included in the ANOVA were: model size, prompt design style, data type (numeric, binary, or categorical), and IRR-F_1_ (the latter two capturing data ambiguity). Data ambiguity metrics reflecting development and test set were used. Statistical analysis was performed in RStudio (2023.06.1), R (4.3.1) and visualizations were performed in ggplot2 (3.5.1).^40,41^

### Evaluation of LLM-extracted events against structured diagnosis dates

The best-performing Llama workflow for three data fields (*MM type, transplant status*, and *extramedullary disease*) was deployed on routine clinical notes within 120 days of MM diagnosis date as determined by ICD codes. Extracted labels for 200 randomly selected patients with routine clinical documents were plotted on the timelines to evaluate frequency, distribution, and timing of the label occurrences around the structured diagnosis date.

## Results

### Reference datasets for performance evaluation

The test dataset of 125 notes (median length of 977 words, IQR 576-289), representing 125 unique patients with MM, was annotated for the selected data fields (100 notes annotated for all data fields, 25 notes for first-line therapy related data fields: *FLT regimen, FLT response, FLT response date*. Very few notes (<10%) contained information for *FLT response date*; this data field was excluded from further analysis.

For the 12 data fields analyzed, the average Krippendorff’s α was α=0.77 for test and development set (0.83 for test set only). Average IRR-F_1_ was F_1_=0.74 for test and development set (0.79 for test set only). Values by data field are shown in Supplementary Table 2.

### Comparison of workflow performance

In total, five LLM workflows (four Llama-based, and one BERT-based) were deployed. Performance of each workflow is shown in Figure 1a. Spearman’s rank correlation coefficients were calculated for continuous data fields (*ECOG score, Plasmacytosis percentage, MM diagnosis date*, and *FLR start date*) (see Figure 1b). Sensitivity analysis for the best-performing Llama workflow by data field is available in Supplementary Note 5.

**Figure 1:**
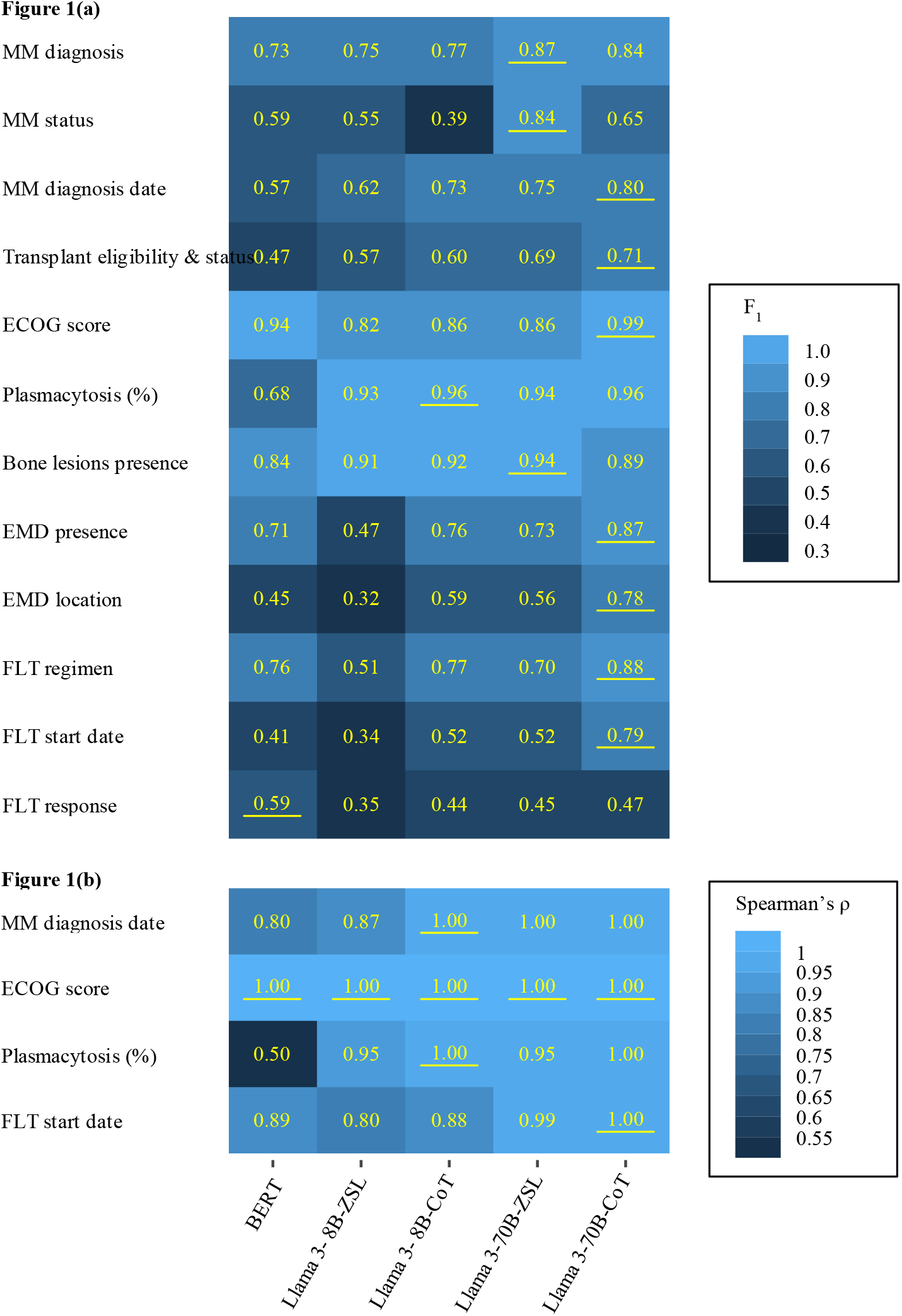
**(b)**;F_1_ score by workflow and data field; **(b)**: Spearman’s rho for numeric data fields.

F_1_ statistics ranged widely by workflow and data field (F_1_ = 0.32-0.99). Llama 70B-CoT had the highest performance on seven out of 12 data fields overall. Notably, the BERT workflow outperformed Llama 8B with both prompt types for three fields, and outperformed 8B-ZSL in four additional fields. For data fields with continuous values, Spearman rank correlations of predictions with the reference data showed mostly near-perfect prediction by 70B workflows.

Figure 2 demonstrates the F_1_ statistics distribution for each Llama workflow compared to BERT workflow. Larger model size and CoT prompting improves performance compared to BERT workflow more consistently

**Figure 2.**
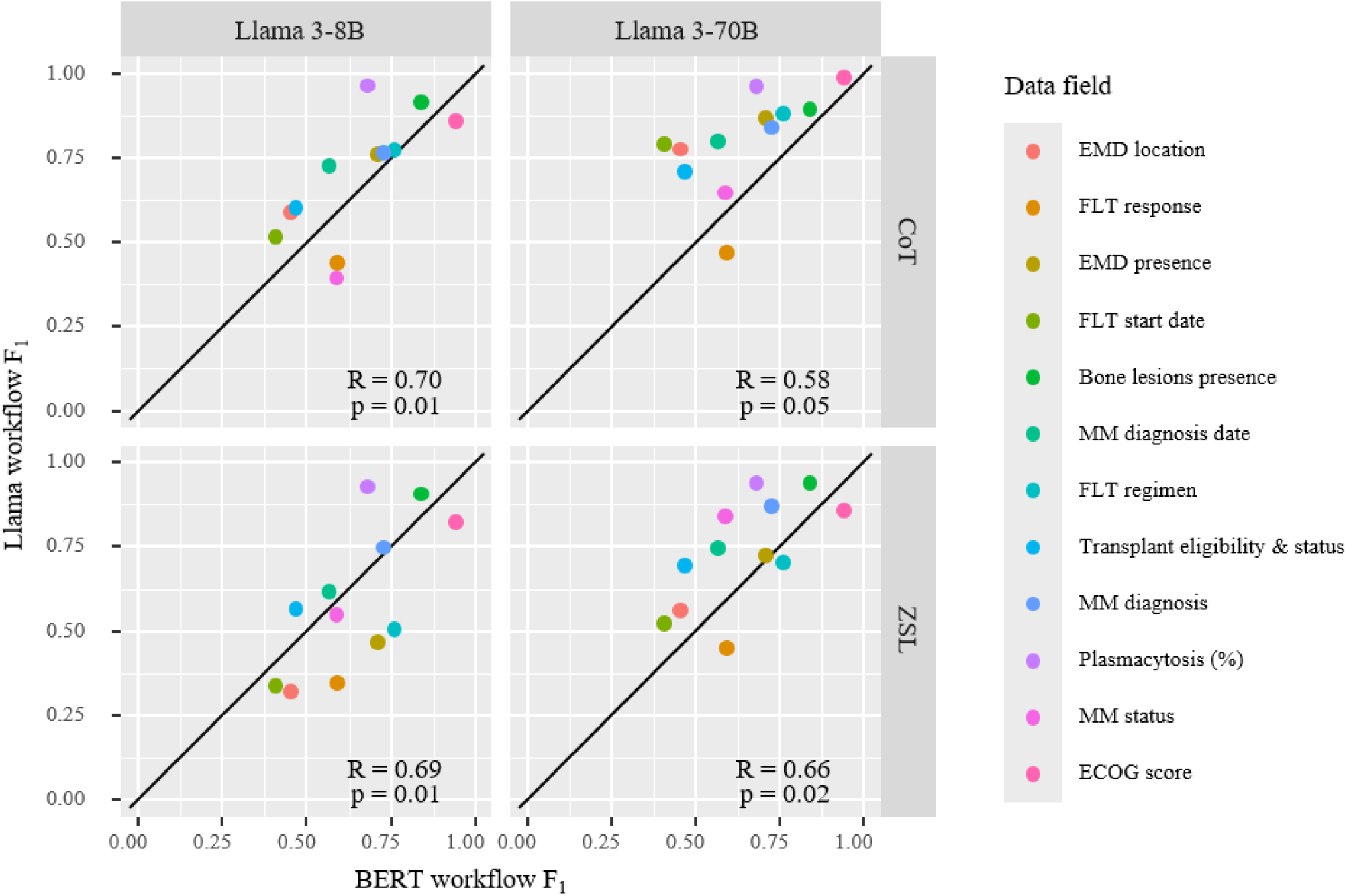
Llama vs BERT workflow F_1_ by data field. R: Pearson correlation coefficient, p: p-value.

### Drivers of Llama workflow performance

ANOVA and pairwise analysis (see Table 2 and Table 3) were used to quantify the impact of model size, prompting technique, and data field characteristics (type and IRR-F_1_) on Llama workflow F_1_ statistics.

**Table 2.** ANOVA test for predictors of Llama workflow F_1_.

| Table 2. ANOVA test for predictors of Llama workflow $F_1$ | | | |
| --- | --- | --- | --- |
| Variable | Statistic | p-value | Significance |
| Model size | 11.78 | $1.36 \times 10^{-3}$ | ** |
| Prompt style | 4.97 | $3.12 \times 10^{-2}$ | * |
| IRR-$F_1$ | 21.70 | $3.20 \times 10^{-5}$ | *** |
| Data field type | 10.92 | $1.52 \times 10^{-4}$ | *** |

**Table 3a.** Univariate testing of predictors of Llama workflow F_1_.

| Table 3a. Univariate testing of predictors of Llama workflow F<sub>1</sub> |  |  |  |  |  |
| --- | --- | --- | --- | --- | --- |
| Variable | Estimate | Statistic | p-value | Significance | Test |
| Model size (70B vs 8B) | 0.13 | 6.79 | $6.37 \times 10^{-7}$ | *** | Paired t-test |
| Prompt style (CoT vs ZSL) | 0.08 | 3.14 | $4.57 \times 10^{-3}$ | ** | Paired t-test |
| IRR-F<sub>1</sub> | 1.02 | 3.52 | $9.87 \times 10^{-4}$ | ** | OLS |
| Data field type | (See pairwise below) | 5.09 | $1.01 \times 10^{-2}$ | * | ANOVA |

**Table 3b.** Pairwise comparison of data field type on Llama workflow F_1_.

| Table 3b. Pairwise comparison of data field type on Llama workflow F<sub>1</sub> |  |  |  |  |  |
| --- | --- | --- | --- | --- | --- |
| Type | Estimate | Statistic | p-value | Significance | Test |
| Binary - categorical | 0.18 | 2.57 | 0.01 | * | Paired t-test |
| Binary - numeric | 0.04 | 0.48 | 0.64 |  | Paired t-test |
| Categorical - numeric | −0.15 | −2.61 | 0.01 | * | Paired t-test |

All variables significantly impact Llama workflow performance (ANOVA *p* < 0.05). IRR-F_1_, capturing data field ambiguity, has the largest impact on workflow F_1._ Increasing model size (from Llama-8B to Llama-70B) has twice the impact on improving F_1_ as using CoT prompting instead of ZSL.

### Comparing NLP workflow performance by inter-rater reliability score

As shown in Figure 3, IRR-F_1_ varied across data fields from 0.61 to 0.88. For all data fields, a larger model size consistently improved LLM workflow performance, although the improvement magnitude was generally higher for more ambiguous data fields (lower IRR-F_1_). Response to CoT prompting was heterogeneous: three data fields (*Bone lesion presence, MM diagnosis, and MM status*) experienced worse performance with CoT than with ZSL; *Plasmacytosis percentage* was essentially equivalent between the two methods. Worse performance with CoT prompting occurred more often in data fields demonstrating low ambiguity (high IRR-F_1_, greater than 0.75). More ambiguous data fields (low IRR-F_1_, under 0.75) were substantially helped by CoT prompting.

**Figure 3.**
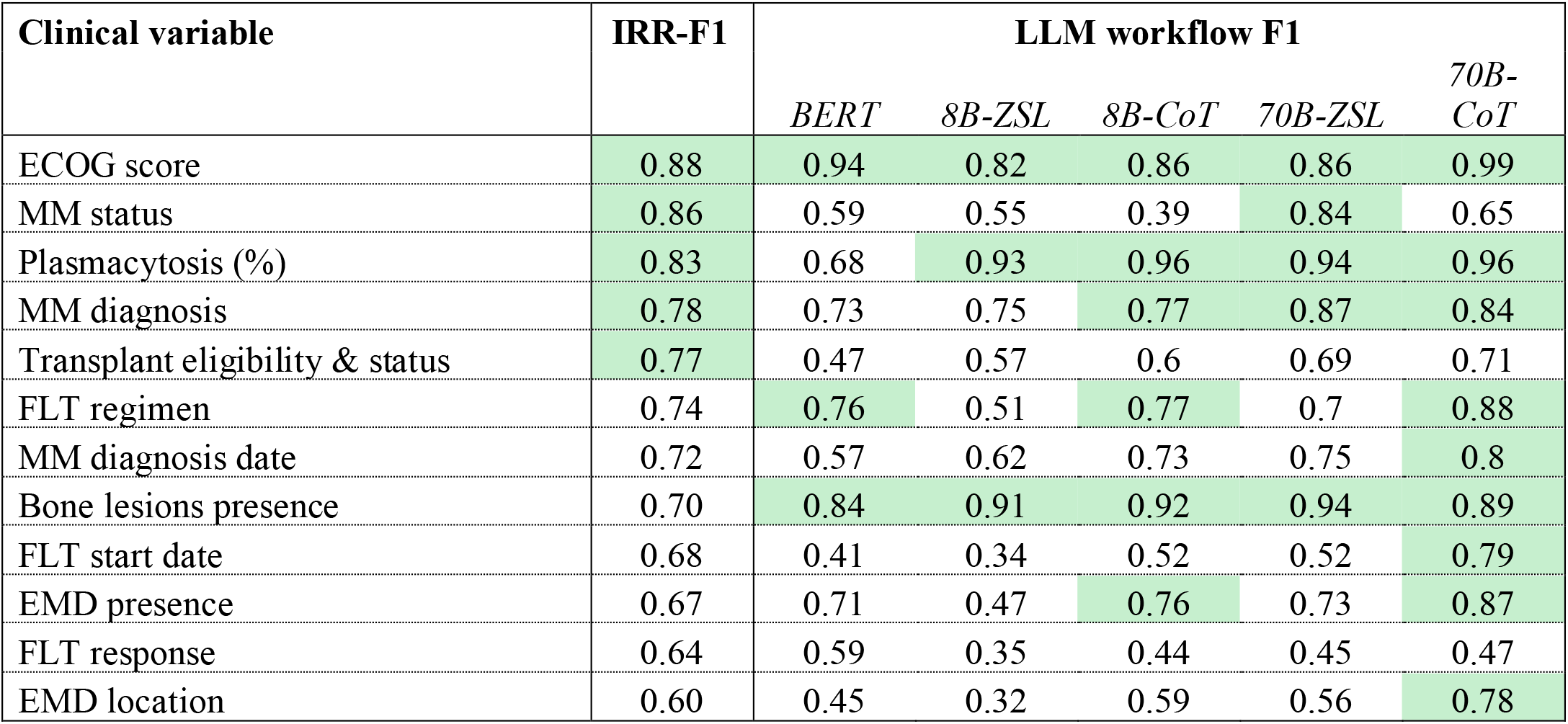
LLM workflow F_1_ in comparison to inter-rater agreement measured by IRR-F_1_. Green-highlighted cells indicate F_1_ statistic over 0.75.

### Exploratory analysis of LLM-extracted events timing vs. structured diagnosis date

Labels for *MM status, Transplant eligibility & status*, and *EMD presence* extracted by the top-performing LLM in a 200-patient subcohort were used to create patient timelines. Figure 4a shows patient-level timelines for 20 illustrative patients (to protect patient-level data), demonstrating the recurrence of labels in a ±120-day window around the MM diagnosis date. Figure 4b shows the aggregated distribution of first label occurrence in each data field class for the entire 200-patient cohort. The most frequent labels occur within a narrow window around the diagnosis date. “Newly diagnosed” and “EMD present” labels cluster most closely to the structured diagnosis date, versus other labels which show more dispersed incidence. Review by authors with medical training (ASB, AOD, AK, MM, SKR, GV) concluded that these distributions were clinically reasonable based on timing of diagnostic testing and documentation.

**Figure 4.**
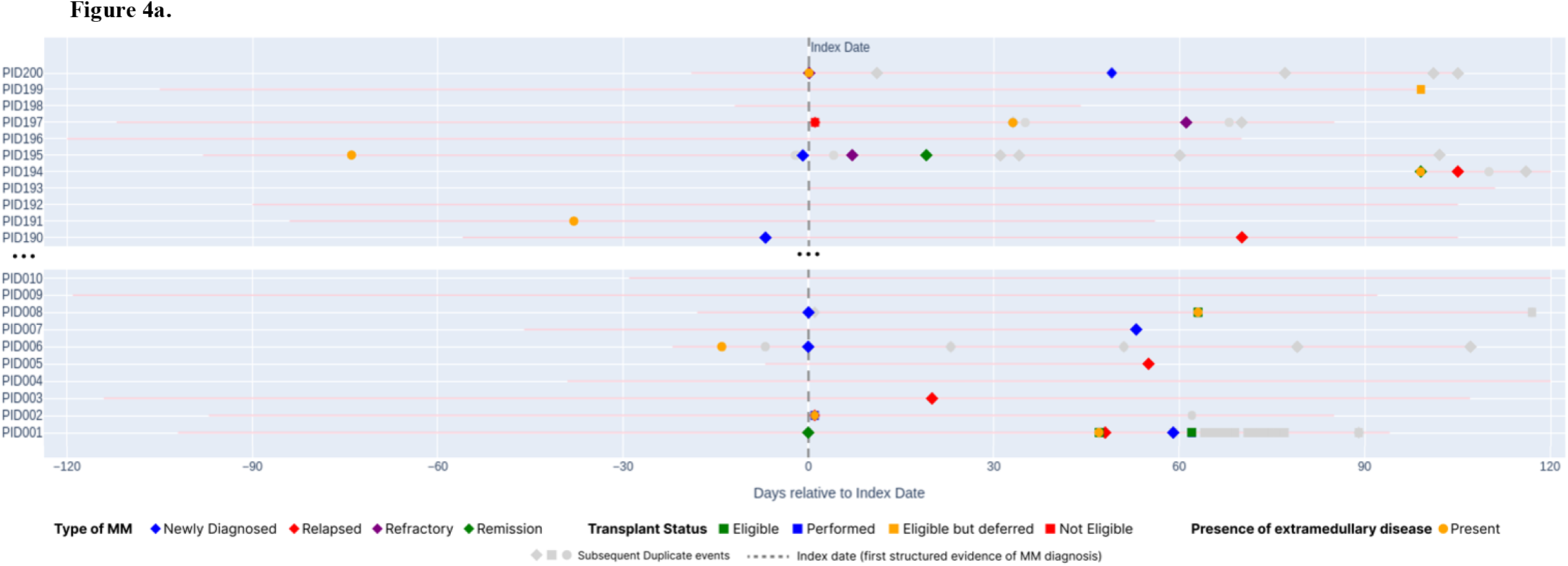
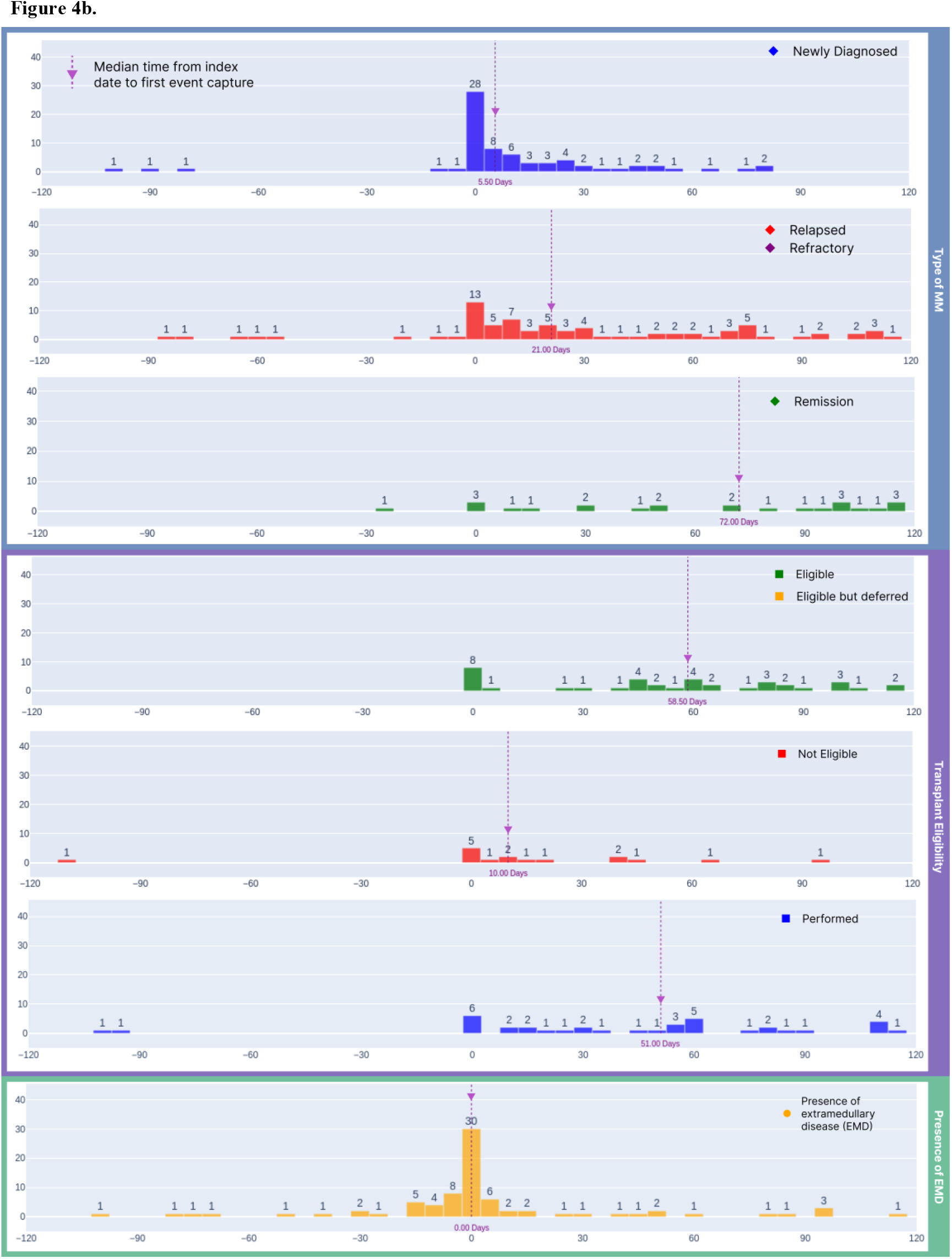
**(a)** Illustrative synthetic patient-level timelines, synthesized using the event rates observed in the 200-patient sample, show extracted labels categorized by type of MM (diamonds), transplant eligibility (squares), and EMD presence (circles), plotted by days relative to the index date (vertical dashed line). First occurrences are colored, and subsequent duplicates are grey. (b) Histograms summarize the timing of first label occurrence, highlighting median extraction times and clustering patterns around the index date.

## Discussion

To accelerate RWD generation, time and resource efficiency in information extraction from unstructured text is paramount. LLMs are increasingly used to improve clinical IE efficiency but must be deployed judiciously.

We tested five NLP workflows for clinical IE to understand performance variability and infer appropriate selection of NLP technique, model size, and prompt design. Our workflows show variability across 12 data fields (F_1_ = 0.59—0.99 for the best NLP workflow). Subsequent statistical analysis of Llama workflow indicated that inter-rater reliability, model size, and prompt design were all significantly associated with performance (ANOVA *p* < 0.05).

All abstraction techniques, both human and machine, are challenged by data ambiguity. Even rigorously trained human abstractors err at a rate of 1-2%; to correct for this, double abstraction with expert arbitration is the gold standard.^42^ We use this human error rate, quantified as IRR-F_1_, as a surrogate for data ambiguity in analyzing Llama workflow performance. IRR-F_1_ was significantly associated with Llama workflow performance (ANOVA *p* = 3.20 × 10^−5^): greater human accuracy predicted higher performance with a point-for-point improvement in F_1_ statistic (OLS *p* = 9.87 x 10^-4^). Since reference dataset creation is a standard NLP workflow development step, IRR-F_1_ can be calculated readily. We suggest IRR-F_1_ as a novel guide for selecting IE approach.

Our data shows that IRR-F_1_ of 0.75 is a reasonable threshold for flagging a challenging data field. Challenging fields, with IRR-F_1_ under 0.75, tend to see a greater performance gain in F_1_ when a larger LLM and CoT prompting is used, compared to the impact seen in simple data fields. In simple fields, with IRR-F_1_ over 0.75, NLP workflow performance is typically higher and there is less improvement with more powerful IE approaches. By using this rule-of-thumb, researchers can increase efficiency in RWD generation without sacrificing data quality.

As expected, larger LLM size is significantly associated with higher F_1_ statistics (ANOVA *p* = 1.36 × 10^−3^). Llama 3-70B performs better and more consistently across data fields than Llama 3-8B. The impact is smaller for simple fields, especially numeric or binary ones, which were satisfactorily extracted by Llama 3-8B and BERT workflows. Larger LLMs, with higher resource requirements, can be reserved for challenging fields where their value will be most realized. Data field type indicated categorical variables were significantly more challenging for NLP models (ANOVA *p* = 1.52 x 10^-4^ for *type* variable; p = 0.01 for pairwise comparison of categorical versus binary or numeric), thus categorical simple fields merit second priority for resource intensive approaches. Researchers without access to a larger model may want to consider model fine-tuning or manual abstraction for challenging fields.

While CoT prompt design was expected to improve LLM performance, study results indicate more nuanced application of CoT prompting for best results. Challenging fields experience significant improvement with CoT prompting (ANOVA *p* = 3.12 × 10^−2^), which should be the preferred approach. Simple fields generally show minimal improvement with CoT prompting and occasional worse results which we hypothesize is due to LLM “over-thinking”. We recommend using ZSL prompting as a default in simple fields, with selective exploration of where CoT might improve results.

Figure 5 summarizes our recommendation for prioritizing resources for data field extraction.

**Figure 5.**
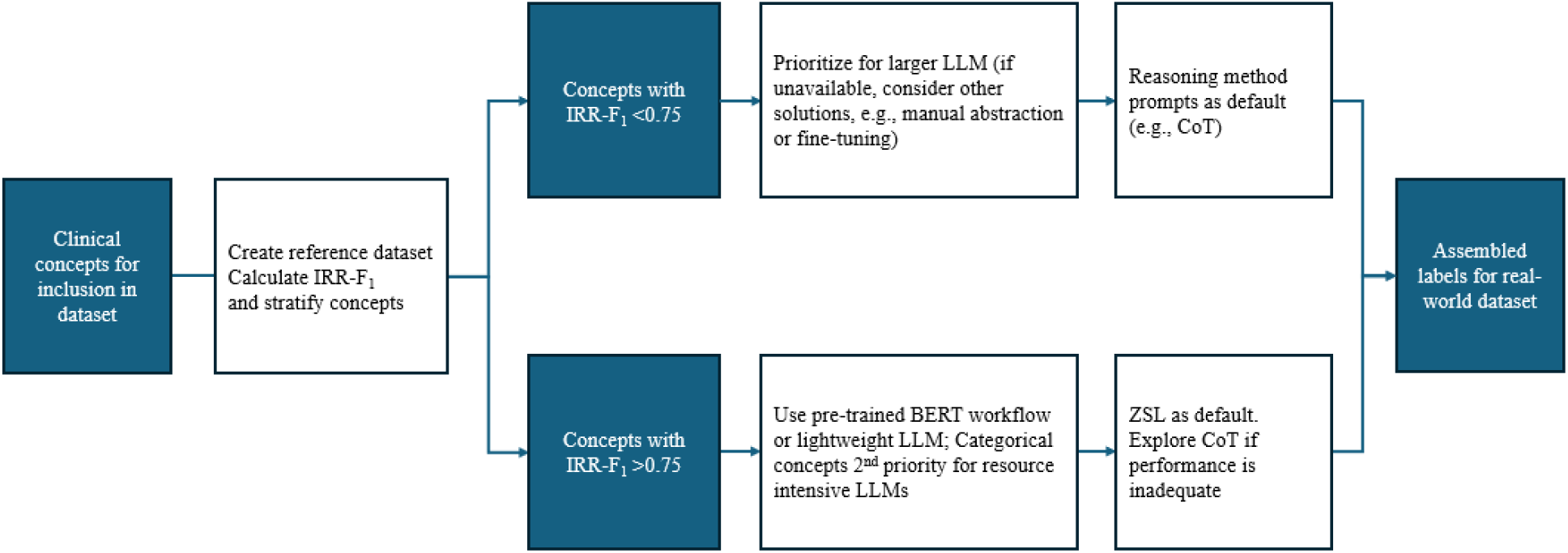
Evaluation criteria for designing IE workflow by data field.

Finally, we constructed patient journey timelines for clinical concepts of interest. We demonstrate that patient journey timelines can provide a sense check for IE performance. Furthermore, the timelines reveal label redundancy in patient charts. Clinical documentation is inherently incomplete, and clinical characteristics evolve along the clinical journey, e.g., a radiology report may not mention transplant status; disease status may change from newly diagnosed to relapsed. NLP tool optimization will not surmount information missingness from a single note or changes over time. Patient journey timelines with repetition of data labels could help solve the uncertainty or incompleteness of any single note when generating labels for a real-world data set. Deploying NLP models over a patient record, rather than a single note, and deriving a consensus label within a timeframe could produce more accurate labelling. Development of a logical method to using information repetition demonstrated along the patient journey to create higher-accuracy real-world datasets would extend this research.

This analysis has two limitations inherent to LLM utilization. LLM performance is highly sensitive to prompt design, which is difficult to quantify, and to model pre-training, which here is limited to the corpora used for Llama models. We could not exhaustively test all models or potential prompts; demonstrated applications of this proposed approach using different NLP models and prompts will support generalization.

## Conclusion

This study analyzes the performance of various NLP models (BERT-based and foundation open-source LLMs of different sizes) in extracting data fields relevant to MM for RWD generation. We show that inter-rater reliability is the largest driver of NLP performance. We propose using an inter-rater reliability metric as a novel guide to efficiently approach information extraction for real-world dataset creation for time and resource allocation without sacrificing data quality.

## Authors

ASB, AOD, DL, MM, GV, AJV, and JG were instrumental in the conception and design of the research. SKR and GV developed the abstraction guidelines and trained the abstractors, with ASB and AK reviewing the guidelines. SKR served as the lead abstractor and played a key role in ground truth dataset generation. MM designed and conducted the BERT-based information extraction, while AA and GV designed and conducted the LLM-based information extraction. DL and AOD performed the formal statistical analysis of the results. ASB and AJV provided high-level guidance, supervision, and resources. ASB, AOD, AK, MM, GV, and SKR contributed to the exploratory analysis and interpretation of patient record-level LLM-based extractions. DL and GV created the figures. AOD, GV, and MM drafted the manuscript, with ASB, AJV, DL, AA, JG, and AK critically revising it. All authors read and approved the final manuscript and supplementary materials. AOD, DL, and GV are co-first authors, and ASB and AJV are co-corresponding authors.

## Acknowledgements

The authors thank Akash Anand (nference) for contributing to the study design and development of BERT and LLM workflows; Praveen Kumar M (nference) for contributing to development of the abstraction guidelines and interpretation of patient record-level LLM-based extractions and manuscript review; Purushotham Sinha (nference) for contributing to setting up the LLM private inference infrastructure and supplementing the analysis; Sai Hanitha and Poorvika Babu (both of nference) for contributing as independent abstractors for dataset creation; Ajit V Rajasekharan (nference); Tommaso Mansi (Johnson & Johnson) for reviewing the manuscript; and Venky Soundarajan (nference) for providing high-level guidance, supervision, and resources.

## Data availability

This study involves the analysis of de-identified Electronic Health Record (EHR) data via the nference Analytics Platform. The data shown and reported in this manuscript was extracted from this environment using an established protocol for data extraction, aimed at preserving patient privacy. The data has been de-identified pursuant to an expert determination in accordance with the HIPAA Privacy Rule. Any data beyond what is reported in the manuscript, including but not limited to the raw EHR data, cannot be shared or released due to the parameters of the expert determination to maintain data de-identification. For additional details regarding the nference Analytics Platform, please contact the corresponding authors.

## Code availability

Annotation protocol, Llama workflow prompts, and BERT workflow rules and model performance metrics are available in Supplementary Materials.

## Supplementary materials

Supplementary Note 1: Data selection and evaluation metrics

Supplementary Note 2: Abstraction protocol and metrics

Supplementary Note 3: Llama workflow prompts

Supplementary Note 4: BERT workflow rules and model performance metrics

Supplementary Note 5: Llama workflow sensitivity analysis

