## Supplementary Note 1 for "Automated abstraction of clinical parameters of multiple myeloma from real-world clinical notes using large language models"

### Supplementary Note 1: Data Selection & Evaluation Metrics

#### 1.1 Data selection

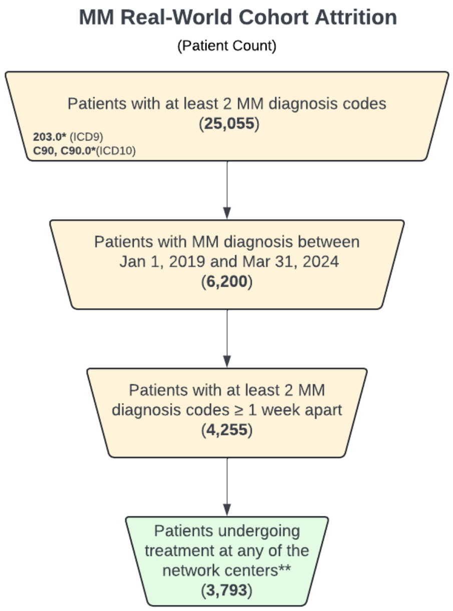


**Supplementary Figure 1.1a.** Cohort attrition diagram: Structured codes 203.0*(ICD-9), and C90, C90.0* (ICD-10) were used for multiple myeloma; * indicates all the children codes within the parent code. ** criteria for determining treatment were assumed if patient had at least one hospital encounter within six months of the diagnosis date and another beyond six months.

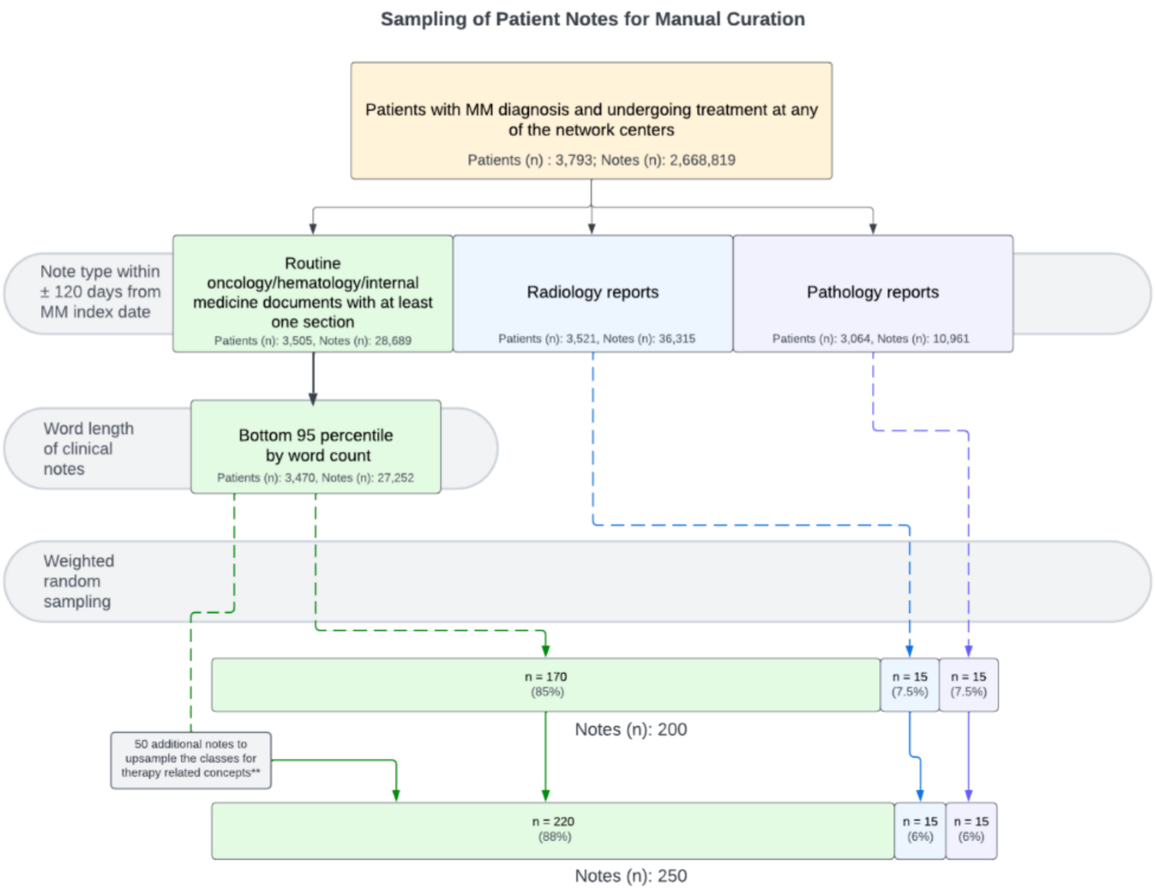


**Supplementary Figure 1.1b.** Sampling and note extraction pipeline:
***Therapy-related data fields include first-line therapy (FLT), date of start of FLT, response to FLT and date of recorded response to FLT.*

Notes within 120 days of diagnosis date and either (1) routine clinical notes from internal medicine, oncology and hematology services; (2) radiology reports; and (3) pathology reports were retained. The longest 5% were excluded because they challenged the optimal context length of the LLMs. On review, the main driver of high word count was duplicative text, likely due to copy-pasting from prior notes. 200 notes were selected using weighted random sampling proportional to frequency of note type.

- 85% notes from internal medicine, oncology, or hematology practice
  - 40% sampled on diagnosis date
  - 40% furthest note within ±120 days of the diagnosis date
  - 20% elsewhere within ±120 days of the diagnosis date
- 7.5% from Radiology practice within ±120 days of the diagnosis date
- 7.5% from Pathology practice within ±120 days of the diagnosis date

50 additional notes were selected because therapy-related data fields (first-line therapy (FLT), date of start of FLT, response to FLT and date of recorded response to FLT) had low coverage after abstraction. The total sampled notes represent 250 unique patients and were divided into development and test sets of 125 notes each.

#### 1.2 Evaluation Metrics

Macro-$F_1$ scores were used to evaluate the performance of the NLP workflows. For multi-label fields such as dates, macro-$F_1$ score was substituted with a weighted (task-specific) $F_1$. Data fields with continuous values were assessed using Spearman's rank correlation coefficient (Spearman's $\rho$).

**Supplementary Table 1.2a:** Weighted (task-specific) $F_1$ metric definitions

| **Condition** | **Ground Truth (GT)** | **Prediction** | **Classification Outcome** |
|---|---|---|---|
| Ground truth and prediction both present and match | Captured | Captured and matches ground truth | True Positive (TP) |
| Ground truth and prediction both absent | Not captured | Not captured | True Negative (TN) |
| Ground truth absent but prediction present | Not captured | Captured | False Positive (FP) |
| Ground truth present but prediction absent | Captured | Not captured | False Negative (FN) |
| Ground truth present and prediction present, not matched | Captured | Captured but does not match | False Positive (FP) |

**Supplementary Table 1.2b:** Data fields and evaluation metric used.

| Concept | Data field | Value type and range | Evaluation metric |
|---|---|---|---|
| MM disease state | *1. MM diagnosis* | Categorical *Yes / Likely / No / Unspecified** | Macro $F_1$ |
| | *2. MM status* | Categorical<br>*Newly diagnosed / Relapsed / Refractory / Remission* | Macro $F_1$ |
| | *3. MM diagnosis date* | Datetime<br>*YYYY-MM-DD* | Task-specific $F_1$ score, Spearman's $\rho$ |
| Transplant eligibility & status | *4. Transplant eligibility & status* | Categorical<br>*Eligible / Eligible but deferred / Ineligible / Performed / Unspecified** | Macro $F_1$ |
| Eastern Cooperative Oncology Group (ECOG) performance status scale | *5. ECOG score* | Integer<br>*0—5* | Macro $F_1$, Spearman's $\rho$ |
| Plasmacytosis | *6. Plasmacytosis percentage* | Integer<br>*0-100%* | Macro $F_1$, Spearman's $\rho$ |
| MM-related bone lesions | *7. MM-related bone lesion presence* | Binary<br>*Yes / No / Unrelated / Unspecified** | Macro $F_1$ |
| Extra-medullary disease (EMD) | *8. EMD presence* | Binary<br>*Present / No / Unspecified** | Macro $F_1$ |
| | *9. EMD location* | Binary<br>*Paramedullary / Soft tissue / Unspecified** | Macro $F_1$ |
| HCP-documented First line therapy (FLT) | *10. FLT regimen* | Categorical<br>*Set of drugs and procedures, mapped to NDC or RxNorm and HCPCS or CPT*** | Task-specific $F_1$ score |
| | *11. FLT start date* | Datetime<br>*YYYY-MM-DD* | Task-specific $F_1$ score, Spearman's $\rho$ |
| HCP-documented FLT response | *12. FLT response* | Categorical<br>*IMWG 2016 response categories: Stringent Complete Response / Complete Response / Very Good Partial Response / Partial Response / Minimal Response / No Response / Progressive Disease / Unspecified** | Macro $F_1$ |
| | *13. FLT response date* | Datetime<br>*Formatted as YYYY-MM-DD* | Task-specific $F_1$ score, Spearman's $\rho$ |
