## Supplementary Note 2 for "Automated abstraction of clinical parameters of multiple myeloma from real-world clinical notes using large language models"

### Supplementary Note 2: Abstraction Protocol and Statistics

#### 2.1 Abstractors

The abstractors were clinicians with 2-3 years of experience in primary and intensive care settings, Abstractors underwent structured abstraction training to ensure consistency and accuracy in data extraction during a pilot abstraction phase on a separate MM patient notes dataset. The abstraction protocol was refined under the oversight of oncology experts to ensure clarity and maintain objectivity.

#### 2.2 Multiple Myeloma (MM) Tagging Guidelines

This document provides guidelines for annotators reviewing clinical notes related to multiple myeloma. Annotators are expected to thoroughly read the clinical notes and answer a series of questions based on the content of these notes.

#### General Guidelines

1. **Inference and Corroboration:**
    - Make inferences from all available context **within** the note.
    - Cross-corroborate the following specifically:
        - Attributability of bone lesions with Multiple Myeloma (MM)
        - Presence and location of extramedullary disease
        - State of MM disease
        - Response to the first line of treatment
        - Bone Marrow Transplant Eligibility
    - Ignore the index date, PID, MM_index_date, and Note_date at the beginning of the note for inference. They are for reference only.

2. **Annotation:**
    - Annotate all relevant context for each question.
    - For date-related questions, use the following interpretations:
        - On: The diagnosis/regimen/response occurred on the mentioned date.
        - Before: The diagnosis/regimen/response occurred before the mentioned date.
    - Annotate complete sentences containing relevant tokens, not just the tokens themselves. Capture from the start to the terminating punctuation or line breaks.
    - Split the annotation span appropriately if multiple contexts from the same sentence answer different questions.
    - For questions relating to drugs or regimens only annotate the drug/regimen terms and not the whole sentence.

3. **Tokens:**
    - Ensure all relevant tokens are entered to be highlighted as part of the note:

myeloma, bone, marrow, osteolytic lesions, skeletal lesions, diagnos, regimen, treatment, chemotherapy, therapy, management, bortezomib, velcade, daratumumab, darzalex, lenalidomide, revlimid, dexamethasone, decadron, thalidomide, thalomid, cyclophosphamide, cytoxan, neosar, line, treatment line, therapy line, complete, partial, response, melphalan, alkeran, prednisone, relapse, refractory, relapsed refractory, carfilzomib, kyprolis, ixazomib, ninlaro, elotuzumab, empliciti, isatuximab, sarclisa, selinexor, xeloda, doxorubicin, adriamycin, doxil, etoposide, vp-16, cisplatin, platinol, pomalidomide, pomalyst, revlimid, revlemid, extramedullary, extramedullary plasmacytoma, ecog, eastern cooperative oncology group, transplant, stem cell transplant, hsct, asct, plasmacytoma, extramedullary plasmacytoma, paramedullary, soft tissue, plasma, cr, complete response, vgpr, very good partial response, pr, partial response, pd, progressive disease, progressive, progression, regim, regimen, date, vrd, vcd, vd, vmp, krd, drd, d-vrd, d-vcd, d-vd, d-vmp, d-krd, d-drd, dara-vrd, dara-vcd, dara-vd, dara-vmp, dara-krd, dara-drd, dara, vr, vd, v-rd, v-cd, v-md, d-vr, d-vc, d-vmp, d-krd, d-drd

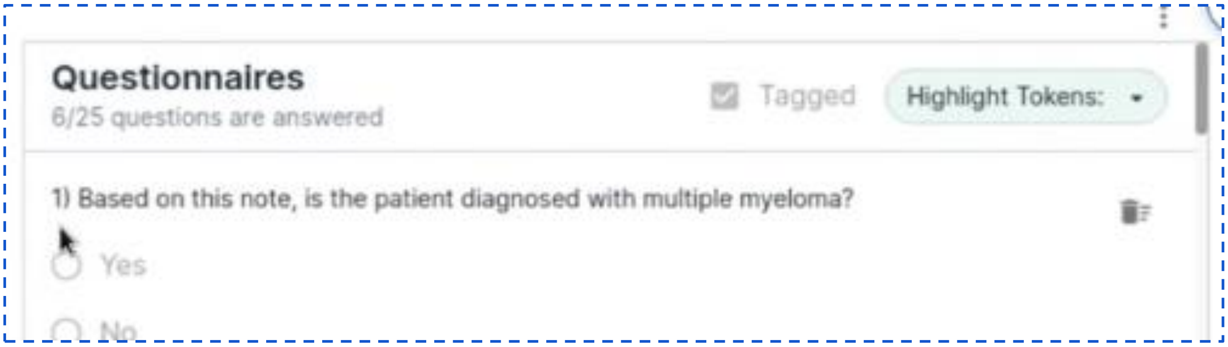


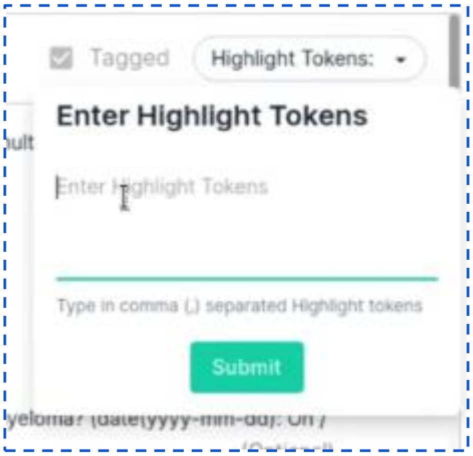


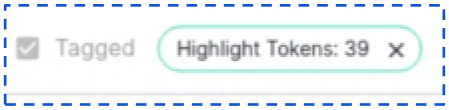


### Questionnaire Guidelines

#### Diagnosis and State of Disease

1. **Based on this note, is the patient diagnosed with multiple myeloma?**
   - **Yes:** Definitive diagnosis of MM.
   - **No:** Conditions like smoldering myeloma, Plasma cell leukemia, MGUS(Monoclonal gammopathy of undetermined Significance), solitary plasmacytoma, POEMS syndrome, Systemic AL amyloidosis.
   - **Likely:** Related plasma cell dyscrasias or MM is mentioned as a differential.
   - **Unknown:** No mention of MM.

2. **What is the earliest date of diagnosis of multiple myeloma?**
   - Insert the date **<yyyy-mm-dd>**: On / Before.
   - If only month and year are mentioned, consider the first day of the month.

    - If only year is mentioned then consider the note’s month as the reference and the first day of the month.

3. **For the above what is the state of Multiple Myeloma?**
    - **Newly Diagnosed:**
        i. Identify newly diagnosed patients who have received no **prior** therapy for MM (emergency interventions like bisphosphonates or steroids can be excluded from treatment history while assessing for this)
        ii. (OR) Currently on first-line therapy or planning to start first-line treatment. Refer to the appendix for criteria. (Current note is within 3 months of initial diagnosis of multiple myeloma - if available in **note context**)
    - **Remission:**
        i. No sign of active multiple myeloma. or Complete Response (CR) is achieved. *(Refer to the appendix for objective criteria.)*
        ii. When available, check for physician sentiments on serum and urine tests, imaging studies, and bone marrow biopsies to confirm remission status.
    - **Relapse:**
        i. Patients with re-emerging symptoms, increased M-protein levels, new lesions, or other clinical indicators of disease return. Refer to the appendix for criteria. (e.g., old MM, entered remission and then signs of Relapse were discussed)
        ii. An increase in M-protein by 25% from the lowest response level (provided that the absolute increase is at least 0.5 g/dL). Development of new bone lesions or soft tissue plasmacytomas. Return of symptoms like anemia, hypercalcemia, or renal dysfunction.(CRAB)
    - **Refractory:**
        i. Primary Refractory, where patients do not achieve atleast a minimum response(MR) after initial therapy for 6 months or (OR) patients showing progression.
        ii. Relapsed and Refractory: Patients who relapse within 60 days after stopping therapy or who progress while on therapy.

4. **Add any comments you have with regards to Q.No: 1,2,3**
    - Add any inferable information regarding the state of MM (e.g., remission/relapse) and highlight the context.

**Treatment Regimen**

List of first-line drugs: Generic name: Brand Name

- Bortezomib :
- Daratumumab :
- Lenalidomide :
- Dexamethasone :
- Thalidomide :
- Cyclophosphamide :

5. **Has the patient been administered the first line of treatment regimen for multiple myeloma?** (Annotate the therapy regimen in the note)

The first line setting is the therapy given to patients during their **newly diagnosed** period. Drugs given for relapse are not to be considered as first line. Suppose the patient is an old case in remission or relapse and the note mentions drugs given during their initial diagnosis of multiple myeloma. In that case, you can consider them as first-line drugs.

- **Yes:** Annotate all relevant drugs or regimens in first line setting.
- **No:** Indicate if no first-line treatment was administered.

6. **When was the first line of the treatment regimen started?** (Annotate the date in the note)
   - Insert the date **(yyyy-mm-dd)**.
   - If only month and year are mentioned, consider the first day of the month.
   - If only year is mentioned then consider the note's month as the reference and the first day of the month. **Example:** The patient was treated in 2016 for x, y, z drugs. And the given note date is 2019-07-16 then we shall mark 2016-07-01.

7. **Was the first line of the treatment regimen stopped?**
   - **Yes:** If any drug in the combination therapy was stopped, consider it as the end of the first line.
     i. e.g. If VRD is the first line therapy and V has been discontinued consider that as the end of VRD(Velcade, Revlimid, Dexamethasone).
   - **No:** Treatment regimen continues.

8. **When was the first line of the treatment regimen stopped?** (Annotate the date in the note)
   - Insert the date **(yyyy-mm-dd)**.
   - If only month and year are mentioned, consider the first day of the month.
   - If only year is mentioned then consider the note's month as the reference and the first day of the month.

9. **Add any comments you have with regards to Q.No: 5 to 8**
   - Document any relevant information if any drugs in the regimen were stopped. Annotate appropriately.

**Other Treatment Lines**

10. **Was the patient on any other line of treatment regimen other than first line? If yes, mention them along with the dates when it was started.**
    - <yyyy-mm-dd> : <treatment regimen> :(On / before), <yyyy-mm-dd> : <treatment regimen>:(On / before), e.g., 2017 : KPd, 2017 : Kyprolis

**Treatment Response**

11. **Based on this note, what was the response to the first line of treatment regimen?** (Refer to Appendix for IMWG guidelines)
    - If the patient was only on the first line of the treatment regimen and the response to it is given, mark it for both Q.no 11 and 13 as it is the first and the latest line of the treatment regimen for the patient)
    - Consider the context in the entire note and answer the question appropriately. Annotate all the contexts relevant to deciding to answer the question. If an explicit mention of any of the following is identified, then it can be directly annotated. If no explicit mention is present then refer to the guidelines in **Appendix.**
        - Stringent complete response (SCR)
        - Complete response (CR)
        - Very good partial response (VGPR)
        - Partial response (PR)
        - Minimal response (MR)
        - No response (NR)
        - Progressive disease (PD)

12. **Date of observation of response to the first line of treatment regimen?**
    - Insert the date **(yyyy-mm-dd)**
    - If only month and year are mentioned, consider the first day of the month.
    - If only year is mentioned then consider the note's month as the reference and the first day of the month.

13. **Response to the latest treatment regimen?** (if different from the first line)
    - If the patient was only on the first line of the treatment regimen and the response to it is given, mark it for both Q.no 11 and 13 as it is the first and the latest line of the treatment regimen for the patient)
    - Consider the context in the entire note and answer the question appropriately. Annotate all the contexts relevant to deciding to answer the question.
        - Stringent complete response (SCR)
        - Complete response (CR)
        - Very Good Partial Response (VGPR)
        - Partial response (PR)
        - Minimal response (MR)
        - No response (NR) / stable disease
        - Progressive disease (PD)

14. **What is the date of response to the latest treatment regimen?** (Annotate the date in the note)
    - Insert the date **(yyyy-mm-dd)**: On / Before.
    - If only month and year are mentioned, consider the first day of the month.
    - If only year is mentioned then consider the note's month as the reference and the first day of the month.

15. **Add any comments you have with regards to Q.No: 10 to 14**

- Add any relevant information (e.g., plateau, no progression) and annotate the context.

**Current Status and Other Indicators**

16. **What drugs is the patient currently on at this time?**
    - List the drugs/regimens as comma-separated-values, and annotate the context in the note appropriately.

17. **What is the plasmacytosis percentage at this time?** (Capture the latest plasmacytosis percentage)
    - Insert the percentage (e.g., "< 4%").

18. **Is there a presence of any bone lesion related to Multiple Myeloma at this time?**
    - **Yes:** Confirmed MM-related bone lesions.
    - **No:** Absence of MM-related bone lesions or explicitly mentioned unrelated bone lesions.
    - **Others:** Bone lesions without definitive mention of MM relation.

**Extramedullary Disease and Performance Status**

19. **Is there a presence of Extramedullary Disease / Extramedullary Plasmacytoma at this time?**
    1. **Present:** Positive sentiment of disease status
        a. **Direct Evidence**:
            i. Explicit statements indicating the presence of extramedullary disease or plasmacytoma.
            ii. Positive imaging findings or pathology reports confirming extramedullary involvement.
        b. **Inferred Evidence**:
            i. Clinical notes describing symptoms directly related to extramedullary disease (e.g., new lumps, neurological symptoms suggesting spinal cord compression that are not due to primary disease process of MM.).
            ii. Treatment notes mention therapy targeting extramedullary sites (e.g., radiation therapy to a specific soft tissue site).
    2. **Absent:** Negative sentiment of disease status
        a. **Direct Evidence**:
            i. Explicit statements indicating the absence of extramedullary disease.
            ii. Negative imaging findings or pathology reports confirming no extramedullary involvement.
        b. **Inferred Evidence**:
            i. Clinical notes discussing normal physical examinations of areas where extramedullary disease might be suspected.

        ii. Follow-up notes indicating stable disease confined to the bone marrow.
3. **Unknown: Disease mentioned, but sentiment not mentioned**
    a. **Direct Evidence**:
        i. Mention of extramedullary disease without a clear indication of presence or absence.
    b. **Inferred Evidence**:
        i. Notes indicating further evaluation needed (e.g., "extramedullary disease to be ruled out with additional imaging").
        ii. Ambiguous statements in clinical notes that neither confirm nor deny the presence.
4. **Unspecified: No mention of disease status**
    a. **Direct Evidence**:
        i. No reference to extramedullary disease in the note.
    b. **Inferred Evidence**:
        i. Notes discussing other disease aspects without mentioning extramedullary involvement.

**20. Add any comments you have with regards to Q.No: 16 to 19**
- Add any relevant information and annotate appropriately.

**21. What is the site of extramedullary disease?** (Refer to Appendix for definitions on extramedullary disease)
1. **Soft Tissue:**
    a. **Direct Evidence**:
        i. Explicit statements indicating extramedullary disease in soft tissues.
        ii. Imaging findings or pathology reports confirming soft tissue involvement.
    b. **Inferred Evidence**:
        i. Clinical notes describing symptoms consistent with soft tissue involvement (e.g., in muscles, ligaments, organs).
        ii. Notes on physical examinations identifying abnormal soft tissue findings, and physician indicating an association with MM.
2. **Paramedullary:**
    a. Paraspinal region, epidural space, chest wall, and paranasal sinuses. In case of bone involvement, confirm that it is **not** due to the primary disease process of multiple myeloma.
    b. **Direct Evidence**:
        i. Explicit statements indicating para-medullary involvement.
        ii. Imaging findings or pathology reports confirming para-medullary involvement.
    c. **Inferred Evidence**:
        i. Clinical notes describing neurological symptoms consistent with spinal cord or nerve root compression.
        ii. Notes on physical examinations identifying tenderness or abnormalities near the spinal column.

3. **Others**
    a. **Direct Evidence**:
        i. Explicit statements indicating extramedullary disease in other specific organs or tissues.
        ii. Imaging findings or pathology reports confirm the involvement of organs like the liver, kidneys, or lungs.
    b. **Inferred Evidence**:
        i. Clinical notes describing symptoms consistent with specific organ involvement (e.g., jaundice for liver involvement, hematuria for kidney involvement).
        ii. Notes indicating abnormal lab results or physical findings relevant to specific organs.

**22. What is the current ECOG status?**

- Insert the ECOG score (0-4).
- **ECOG Performance Status Criteria:**
    - **0:** Fully active, able to carry on all pre-disease performance without restriction.
    - **1:** Restricted in physically strenuous activity but ambulatory and able to carry out work of a light or sedentary nature (e.g., light housework, office work).
    - **2:** Ambulatory and capable of all self-care but unable to carry out any work activities; up and about more than 50% of waking hours.
    - **3:** Capable of only limited self-care; confined to bed or chair more than 50% of waking hours.
    - **4:** Completely disabled; cannot carry on any self-care; totally confined to bed or chair.
    - **5:** Dead.

**Transplant Status**

**23. Based on the note, what is the transplant status?**

- Performed
    i. Direct mention of the procedure, or “underwent transplant”/ “transplant performed”, “received autologous stem cell transplant” etc.,
- Eligible
    i. Indications that the patient is a suitable candidate for transplant without any deferment or contraindications.
    ii. Look for specific phrases such as “eligible for transplant” or “candidate for stem cell transplant” / “transplant is a viable option”
- Not eligible: Provide the reason.
    i. Indications that the patient is not suitable for transplant, along with a documented reason (if available).
    ii. Specific phrases such as "not eligible for transplant", "contraindicated for transplant", and "transplant not an option due to [reason]".
- Eligible but deferred: Provide the reason.

        i. Cases such as "The patient is eligible for a transplant but it has been deferred due to infection.", "Transplant postponed until patient achieves remission." "Eligible but deferred due to insurance issues." etc.,
    - Not assessed for transplantation. (No clear indication for transplant eligibility/ procedure being performed)

**24. With reference to the question no 23, mention the reason if available**
- Insert the reason.

**25. Add any comments you have with regards to Q.No: 21 to 24**
- Mention the site of extramedullary involvement if the answer to Q21 is "Others".
- Mention other performance indicators like Karnofsky or ADL and annotate appropriately.

#### Guidelines

**Response Criteria (IMWG)**

**<IMWG Uniform Response Criteria | Int'l Myeloma Fn>**

- **sCR:** CR plus normal FLC ratio and absence of clonal cells in bone marrow.
- **CR:** Negative immunofixation on serum and urine, the disappearance of soft tissue plasmacytomas, and < 5% plasma cells in bone marrow.
- **VGPR:** Serum and urine M-protein detectable by immunofixation but not electrophoresis, or > 90% reduction in serum M-protein plus urine M-protein level < 100 mg/24 h.
- **PR:** > 50% reduction of serum M-protein and reduction in 24-hour urinary M-protein by >90% or to < 200 mg/24 h.
- **MR:** NA. (Consider this if no objective measures are present and the physician has mentioned that the patient is tolerating well and **responding** to treatment.)
- **No Response/Stable disease:** Not meeting criteria for CR, VGPR, PR, or PD. (Consider this if the physician has only mentioned about patient tolerating the therapy but have not discussed anything about the response.)
- **Plateau:** NA.
- **Progressive disease:** Increase of > 25% from lowest response value in serum/urine M-component, FLC levels, bone marrow plasma cell percentage, development of new bone lesions or soft tissue plasmacytomas, or hypercalcaemia.
- **Relapse:** Clinical relapse indicators include new soft tissue plasmacytomas, lytic bone lesions, hypercalcemia or reappearance of serum or urine M-protein by immunofixation or electrophoresis or development of >5% plasma cells in bone marrow.

**Extramedullary Disease/ Extramedullary Plasmacytoma:**

**Distinguishing Between Primary and Extramedullary Involvement**

1. **Primary Bone Involvement**:
   - **Osteolytic Lesions**: Multiple myeloma typically causes osteolytic lesions in bones, including the vertebrae and ribs. These lesions are due to the direct infiltration of myeloma cells into the bone marrow and subsequent bone destruction.
   - **Pathological Fractures**: The weakening of bones from osteolytic activity can lead to fractures, which are common in vertebrae and ribs.
   - **Imaging Findings**: Radiographic imaging, including X-rays, CT scans, and MRIs, often shows multiple punched-out lytic lesions or diffuse bone destruction in these areas.
   - **Within Bone Marrow**: Lesions are within the bone marrow of vertebrae or ribs, causing osteolytic destruction.
   - **Common Sites**: Vertebrae and ribs are common sites for primary bone lesions in multiple myeloma due to their rich bone marrow content.
   - **Symptoms**: Bone pain, pathological fractures, and spinal cord compression due to vertebral involvement.
2. **Extramedullary Involvement**:
   - **Definition**: Extramedullary disease refers to the presence of plasma cell tumors outside the bone marrow. This can include soft tissues, organs, and paramedullary sites adjacent to bones but not within the bone marrow itself. This disease process is not a direct result of primary disease process of MM in the adjacent tissues of bone marrow.
   - **Paramedullary Involvement**: Lesions in tissues adjacent to the bones (e.g., epidural space, paraspinal tissues) are considered extramedullary if they do not infiltrate the respective bone marrow.
   - **Outside Bone Marrow**: Lesions are in soft tissues or organs outside the bone marrow. For paramedullary involvement, the lesions are adjacent to but not **within the bone.**
   - **Common Sites**: Soft tissues, liver, spleen, kidneys, lymph nodes, CNS, and paramedullary tissues like the epidural space.
   - **Symptoms**: Can vary widely depending on the location of the extramedullary lesions and may include organ-specific symptoms or neurological deficits if near the spine.

**Example Scenario**

- **Primary Involvement**: A patient with multiple myeloma has osteolytic lesions in the vertebrae seen on MRI, causing vertebral compression fractures and spinal pain. This is due to the primary disease process within the bone marrow.
- **Extramedullary Involvement**: Another patient has a soft tissue mass in the epidural space causing spinal cord compression, with biopsy confirming plasma cells. This is considered paramedullary extramedullary disease.

*****Please adhere to these guidelines meticulously to ensure accurate and consistent annotations.*

#### 2.3 Manual Abstraction Results

**Supplementary Table 2.3a.** Krippendorff's α

| Data field | Krippendorff's α (Development set) | Krippendorff's α (Test set) |
|---|---|---|
| MM diagnosis | 0.7 | 0.9 |
| MM status | 0.8 | 0.9 |
| MM diagnosis date | 0.6 | 0.8 |
| Transplant eligibility & status | 0.5 | 0.8 |
| ECOG score | 0.9 | 0.9 |
| Plasmacytosis percentage | 0.7 | 0.9 |
| MM-related bone lesion presence | 0.7 | 0.9 |
| EMD presence | 0.8 | 0.8 |
| EMD location | 0.6 | 0.6 |
| FLT regimen | 0.8 | 0.9 |
| FLT start date | 0.7 | 0.9 |
| FLT response | 0.7 | 0.7 |
| FLT response date | -0.03 | 0.1 |

**Supplementary Table 2.3b.** Abstractor-Consensus Agreement

$F_1$ scores to measure abstractor-consensus were calculated by comparing the unarbitrated responses of each abstractor with the corresponding arbitrated responses (consensus) for development and test sets.

| **Data field** | **Macro-$F_1$ (Abstractor 1 : Consensus)** | **Macro-$F_1$ (Abstractor 2 : Consensus)** | **Averaged macro-$F_1$ (IRR $F_1$)** |
|---|---|---|---|
| MM diagnosis | 0.78 | 0.78 | 0.78 |
| MM status | 0.89 | 0.83 | 0.86 |
| MM diagnosis date | 0.75 | 0.69 | 0.72 |
| Transplant eligibility & status | 0.83 | 0.72 | 0.78 |
| ECOG score | 0.86 | 0.90 | 0.88 |
| Plasmacytosis percentage | 0.82 | 0.84 | 0.83 |
| MM-related bone lesion presence | 0.70 | 0.70 | 0.70 |
| EMD presence | 0.63 | 0.70 | 0.67 |
| EMD location | 0.59 | 0.62 | 0.61 |
| FLT regimen | 0.82 | 0.67 | 0.75 |
| FLT start date | 0.70 | 0.65 | 0.68 |
| FLT response | 0.71 | 0.56 | 0.64 |
| FLT response date | 0.16 | 0.34 | 0.25 |

### Class distribution

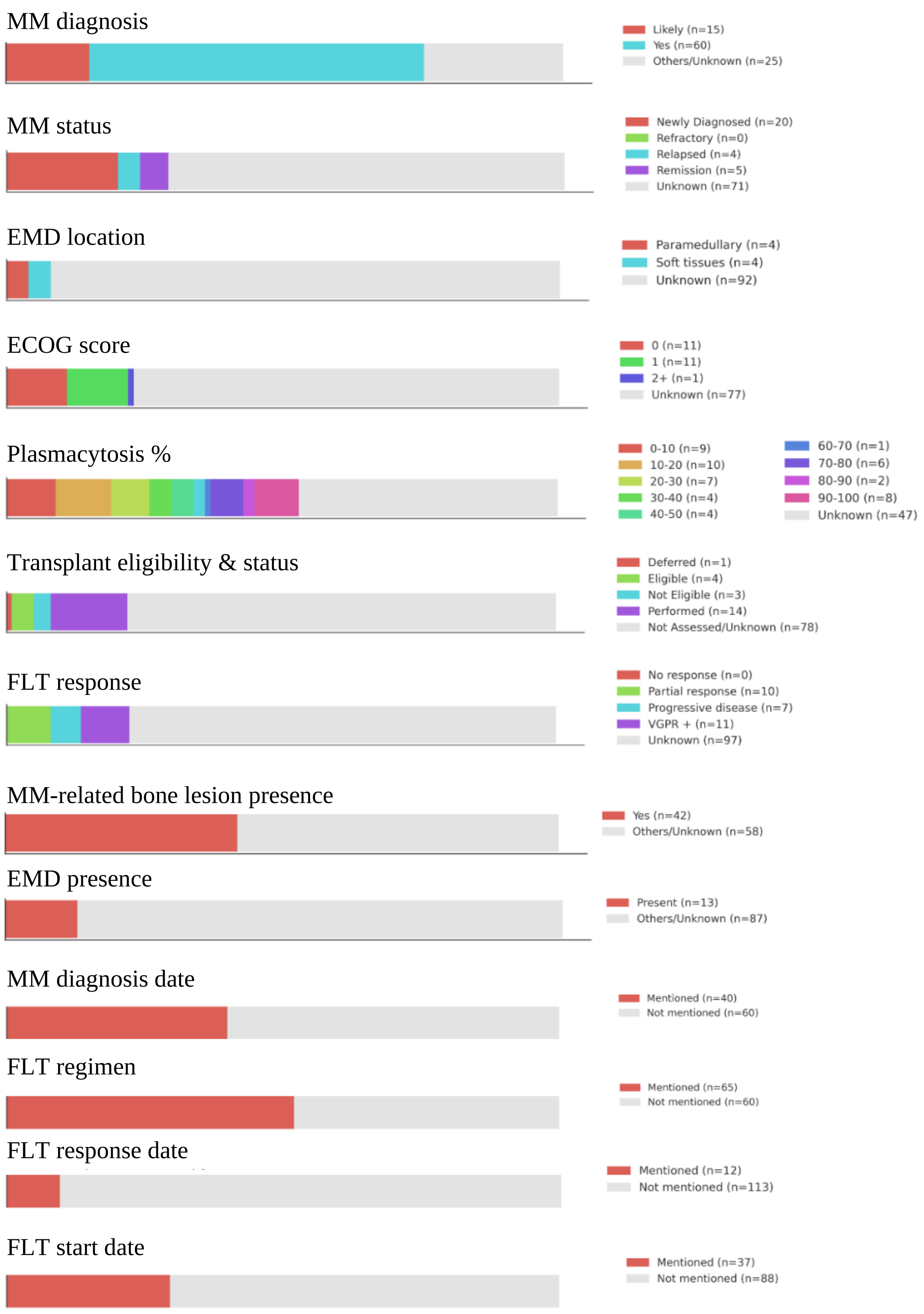


**Supplementary Figure 2.3.** Class distribution by data field
