## Supplementary Note 3 for "Automated abstraction of clinical parameters of multiple myeloma from real-world clinical notes using large language models"

### LLM Prompts

The Chain of Thought (CoT) prompting method uses the `reason` and `answer` keys, while the Zero-Shot (ZSL) method uses the `combined` key from the YAML configuration to construct prompts for each concept. At runtime, `{chunk_text}` is replaced with the patient note, and `validation_pairs` are applied for output standardization when needed.

```yaml
---

bone_lesion:

        reason: |

                Based on the given patient note:

                {chunk_text}

                Has the presence of active bone lesions been confirmed in the patient? If it has then when? Also discuss if the bone lesion related evaluation is in context of Multiple Myeloma

                DO NOT imply or infer anything, only use the information given in the note.

        answer: |

                Based on the reason above answer whether or not the patient has bone lesions related to Multiple Myeloma

                Yes: "if active bone lesion is present and if it's in context of MM diagnosis and evaluation"

                No: "if it's confirmed that there are no active bone lesions present or bone lesions are present but they are not related to MM in any sense"

                NO_INFO: if there was no mention of bone lesions in note

                Provide your answer in following format

                {{"bone_lesions_present": "one of the above labels"}}

                ONLY OUTPUT THE JSON AND NOTHING ELSE NO REASONING NEEDED

        combined: |

                Based on the given patient note:
```

```
                {chunk_text}

                Answer whether or not the patient has bone lesions related to
Multiple Myeloma

                Yes: "if active bone lesion is present and if it's in context
of MM diagnosis and evaluation"

                No: "if it's confirmed that there are no active bone lesions
present or bone lesions are present but they are not related to MM in any
sense"

                NO_INFO: if there was no mention of bone lesions in note.

                Provide your answer in the following format

                {{"bone_lesions_present": "one of the above labels"}}

                ONLY OUTPUT THE JSON AND NOTHING ELSE NO REASONING NEEDED

        validation_pairs:

                bone_lesions_present:

                - 'Yes'

                - 'No'

                - 'NO_INFO'

mm_diagnosis:

        reason: |

                Based on the given patient note:

                {chunk_text}

                Reason about the following questions and then provide an
answer

                1. Diagnosis of the multiple myeloma, i.e., whether or not
the patient has a confirmed diagnosis of MM

                Yes: the patient has MM

                No: the diagnosis of MM was negative for patient

                Likely: if the patient is under evaluation for MM or if the
```

```
doctor has noted possibility of Multiple Myeloma based on some findings

                Precursory: If the patient has some precursory condition like
smoldering MM or MGU and doctor has not considered him for multiple myeloma
evaluation

                NO_MENTION: If note does not contain any information related
to this.

                2. When was it first diagnosed in the patient (Note that,
this patient note was written on {note_date} in case you need to refer to the
date of the note and do not infer from the date of start of treatment or some
lab finding date for the date of diagnosis).

                3. Type of MM(one of NewlyDiagnosed, Refractory, Relapsed and
Remission) - only if mentioned in note otherwise output NO_MENTION

        answer: |

                Based on this can you create a json with following
structure(str, str)

                {{

                "patient_has_mm: one of ('Yes', 'No', 'Likely', 'Precursory'
'NO_MENTION'),

                "date_of_diagnosis": "date in dd-mm-yyyy or dd-mmm-yyyy
format if present in note else 'NO_MENTION'",

                "type_of_mm": one of ('NEWLY_DIAGNOSED', 'REFRACTORY',
'RELAPSED', 'REMISSION', 'NO_MENTION')

                }}

                NOTES:

                For date_of_diagnosis, if day is missing then use 1 in place
of dd, if the month and/or year is missing then pick missing value from the
date when note was written which is {note_date}. DO NOT OUTPUT ANY STRING
OTHER THAN NO_MENTION OR THE DATE IN SPECIFIED FORMAT

                For patient_has_mm: 'Precursory' label should only be used if
Yes, No and Likely is not applicable and the patients has conditions like MGU
or smoldering multiple myeloma.

                ONLY OUTPUT A JSON WITH ABOVE KEYS AND NOTHING ELSE

        combined: |
```

```
                        Based on the given patient note:

                        {chunk_text}

                        Answer following questions?

                        1. Whether or not the patient has a confirmed
diagnosis of MM

                        Yes: the patient has MM

                        No: the diagnosis of MM was negative for patient

                        Likely: if the patient is under evaluation for MM or
if the doctor has noted possibility of Multiple Myeloma based on some
findings

                        Precursory: If the patient has some precursory
condition like smolering MM or MGU and doctor has not considered him for
multiple myeloma evaluation

                        NO_MENTION: If note does not contain any information
related to this.

                        2. When was it first diagnosed in the patient (Note
that, this patient note was written on {note_date} in case you need to refer
to the date of the note and do not infer from the date of start of treatment
or some lab finding date for the date of diagnosis).

                        3. Type of MM(one of NewlyDiagnosed, Refractory,
Relapsed and Remission) - only if mentioned in note otherwise output
NO_MENTION

                        Output a json with following structure(str, str)

                        {{

                        "patient_has_mm": one of ('Yes', 'No', 'Likely',
'Precursory' 'NO_MENTION'),

                        "date_of_diagnosis": "date in dd-mm-yyyy or dd-mmm-
yyyy format if present in note else 'NO_MENTION'",

                        "type_of_mm": one of ('NEWLY_DIAGNOSED',
'REFRACTORY', 'RELAPSED', 'REMISSION', 'NO_MENTION')

                        }}

                NOTES:
```

```
                For date_of_diagnosis, if day is missing then use 1 in place
of dd, if the month and/or year is missing then pick missing value from the
date when note was written which is {note_date}. DO NOT OUTPUT ANY STRING
OTHER THAN NO_MENTION OR THE DATE IN SPECIFIED FORMAT

                For patient_has_mm: 'Precursory' label should only be used if
Yes, No and Likely is not applicable and the patients has conditions like MGU
or smoldering multiple myeloma.

                ONLY OUTPUT A JSON WITH ABOVE KEYS AND NOTHING ELSE NO
REASONING NEEDED

        validation_pairs:

                patient_has_mm:

                - 'Yes'

                - 'No'

                - 'Likely'

                - 'NO_MENTION'

                type_of_mm:

                - 'NEWLY_DIAGNOSED'

                - 'REFRACTORY'

                - 'RELAPSED'

                - 'REMISSION'

                - 'NO_MENTION'

extramedullary_disease:

        reason: |

                Based on the given patient note:

                {chunk_text}

                What can you say about the presence of extramedullary
disease/extramedullary plasmacytoma in the context of multiple myeloma? If
present, discuss the possibility of root cause being the primary disease
process of multiple myeloma? Also comment whether it`s paramedullary or soft
tissue or some other location in the note`s context.
```

```
        answer: |

                Based on this can you create a json with the following
structure:

                {{

                extramedullary_disease: one of ("YES", MAYBE_YES,
"NO_MENTION"),

                location: one of ("soft_tissue", "paramedullary" "other",
"NA")

                }}

                Pick label for extramedullary_disease:

                MAYBE_YES: I. If the presence of em disease is not confirmed
yet

                II. If the paramedullary lesions are identified in the note
but they not directly from the bone-marrow(for example, if the lesions are
found in T12 but there`s no corresponding bone marrow involvement, it will be
labeled as MAYBE_YES)

                YES: If the lesions is confirmed to be extramedullary and is
in the context of multiple myeloma.

                NO_MENTION: If the extramedullary disease is not discussed or
no such lesions are identified

                For location, use soft_tissues or paramedullay depending on
the location of extramedullary disease and if it`s none of these two then use
other. Use NA if extramedullary disease is not present

                ONLY OUTPUT A JSON WITH ABOVE KEYS AND NOTHING ELSE

        combined: |

                Based on the given patient note:

                {chunk_text}

                Output a json with the following structure:

                {{

                extramedullary_disease: one of ("YES", MAYBE_YES,
"NO_MENTION"),
```

```
                location: one of ("soft_tissue", "paramedullary" "other",
"NA")

                }}

                Pick label for extramedullary_disease:

                MAYBE_YES: I. If the presence of em disease is not confirmed
yet

                II. If the paramedullary lesions are identified in the note
but they not directly from the bone-marrow(for example, if the lesions are
found in T12 but there`s no corresponding bone marrow involvement, it will be
labeled as MAYBE_YES)

                YES: If the lesions is confirmed to be extramedullary and is
in the context of multiple myeloma.

                NO_MENTION: If the extramedullary disease is not discussed or
no such lesions are identified

                For location, use soft_tissues or paramedullay depending on
the location of extramedullary disease and if it`s none of these two then use
other. Use NA if extramedullary disease is not present

                ONLY OUTPUT A JSON WITH ABOVE KEYS AND NOTHING ELSE NO
REASONING NEEDED



plasmacytosis:

        reason: |

                Based on the given patient note:

                {chunk_text}

                What can you say about the plasmacytosis percentage in the
patient? If more than one value is present discuss them all with dates when
the evaluation was done

        answer: |

                Based on this can you provide the plasmacytosis percentage
value given for the patient in the following format.

                {{"plasmacytosis_percentage": "plasmacytosis % as given in
the note"}} # if no percentage value is given then the value should be
NO_MENTION
```

```
                If more than one value instance is present, pick latest one

                ONLY OUTPUT THE JSON AND NOTHING ELSE NO REASONING NEEDED

        combined: |

                Based on the given patient note:

                {chunk_text}

                Provide the plasmacytosis percentage value given for the
patient in the following format.

                {{"plasmacytosis_percentage": "plasmacytosis % as given in
the note"}} # if no percentage value is given then the value should be
NO_MENTION

                If more than one value instance is present, pick latest one.
If the latest value present is more than a year old(from when this note was
written) then output NO_MENTION. This note was written on {note_date}.

                ONLY OUTPUT A JSON WITH ABOVE KEYS AND NOTHING ELSE NO
REASONING NEEDED



ecog:

        reason: |

                Based on the given patient note:

                {chunk_text}

                What can you say about the ECOG score(scale 0 to 5) of the
patient?

        answer: |

                Based on this can you provide the ECOG score of the patient
in the following format.

                {{"ECOG_SCORE": "ECOG Score as given in note"}} # if nothing
is mentioned then the value should be NO_MENTION

                ONLY OUTPUT THE JSON AND NOTHING ELSE NO REASONING NEEDED

        combined: |
```

```
                Based on the given patient note:

                {chunk_text}

                Output the ECOG score of the patient in the following format.

                {{"ECOG_SCORE": "ECOG Score as given in note"}} # if nothing
is mentioned then the value should be NO_MENTION

                ONLY OUTPUT THE JSON AND NOTHING ELSE NO REASONING NEEDED



first_line_treatment:

        reason: |

                Based on the given patient note:

                {chunk_text}

                Comment regarding the first line treatment given to patient
for Multiple Myeloma

                Was the first line treatment given? If yes, then comment on
the treatment(/drug/regimen name) that was given, when did it start. Also
what was the response to the treatment and when was the response recorded? If
the date of starting first line treatemnt and response record date is
specifically given then only discuss, don`t imply it using other clues?

        answer: |

                Based on this can you create a json with the following
structure:

                {{

                "first_line_treatment_given": (str) one of ("YES", "NO",
"NO_MENTION"), # Put NO_MENTION if it's not clear from the note

                "treatment": (List[str]) a list of drug names, "if multiple
drugs form a treatment regimen then replace the combination with regimen
names(mapping given below) and append the individual drugs that can't be
merged to the same list. If there's no mention of any first line treatment
drugs, output an empty list."

                "date_started": (str)"date in dd-mm-yyyy or dd-mmm-yyyy
format if present in note else 'NO_MENTION'",

                "treatment_response": (str) "most appropriate response label
```

```
as per exmplanation given in NOTES",

                "treatment_response_record_date": (str) "date in dd-mm-yyyy
or dd-mmm-yyyy format if present in note else 'NO_MENTION'"

                }}

                Only consider following treatment regimens and generic
drugs(if brand name of these generic drugs are given then replace those with
generic names in response).

                {{

                "vrd-lite": [

                "vrd-lite"

                ],

                "vrd": [

                "rvd",

                "vrd",

                "bortezomib-dexamethasone-lenalidomide"

                ],

                "cybord": [

                "cybord",

                "bortezomib-cyclophosphamide-dexamethasone"

                ],

                "krd": [

                "krd",

                "carfilzomib-dexamethasone-lenalidomide"

                ],

                "dara-vrd": [

                "dara-vrd",

                "bortezomib-daratumumab-dexamethasone-lenalidomide"
```

```
],
"rd": [
"dexamethasone-lenalidomide",
"rd"
],
"vd": [
"bortezomib-dexamethasone",
"vd"
],
"dvd": [
"bortezomib-daratumumab-dexamethasone",
"dvd"
],
"dara-cybord": [
"dara-cybord",
"bortezomib-cyclophosphamide-daratumumab-dexamethasone"
],
"vzd": [
"dexamethasone-lenalidomide-zometa"
],
"dara-rd": [
"drd rd",
"daratumumab-dexamethasone-lenalidomide"
],
"vtd": [
```

```
                "vtd",

                "bortezomib-dexamethasone-thalidomide"

                ]

                }}

                NOTES:

                For both the dates, if day is missing then use 1 in place of
dd, if the month and/or year is missing then pick missing value(s) from the
date when note was written which is {note_date} If nothing is clearly
mentioned then output NO_MENTION. DO NOT OUTPUT ANY STRING OTHER THAN
NO_MENTION OR THE DATE IN SPECIFIED FORMAT

                Treatment response labels(Pick most suitable one based on the
IMWG criteria): PARTIAL_RESPONSE, MINIMAL_RESPONSE,
STRINGENT_COMPLETE_RESPONSE, COMPLETE_RESPONSE, VERY_GOOD_PARTIAL_RESPONSE,
NO_RESPONSE, PROGRESSIVE_DISEASE, NO_MENTION # use no mention if there's no
mention of this in the note

                Note that this patient note was written on {note_date} in
case you need to refer to the date of the note.

                ONLY OUTPUT A JSON WITH ABOVE KEYS AND NOTHING ELSE

        combined: |

                Based on the given patient note:

                {chunk_text}

                Can you create a json with the following structure:

                {{

                "first_line_treatment_given": (str) one of ("YES", "NO",
"NO_MENTION"), # Put NO_MENTION if it's not clear from the note

                "treatment": (List[str]) a list of drug names, "if multiple
drugs form a treatment regimen then replace the combination with regimen
names(mapping given below) and append the individual drugs that can't be
merged to the same list. If there's no mention of any first line treatment
drugs, output an empty list."

                "date_started": (str)"date in dd-mm-yyyy or dd-mmm-yyyy
format if present in note else 'NO_MENTION'",
```

```
                "treatment_response": (str) "most appropriate response label
as per exmplanation given in NOTES",

                "treatment_response_record_date": (str) "date in dd-mm-yyyy
or dd-mmm-yyyy format if present in note else 'NO_MENTION'"

                }}

                Only consider following treatment regimens and generic
drugs(if brand name of these generic drugs are given then replace those with
generic names in response).

                {{

                "vrd-lite": [

                "vrd-lite"

                ],

                "vrd": [

                "rvd",

                "vrd",

                "bortezomib-dexamethasone-lenalidomide"

                ],

                "cybord": [

                "cybord",

                "bortezomib-cyclophosphamide-dexamethasone"

                ],

                "krd": [

                "krd",

                "carfilzomib-dexamethasone-lenalidomide"

                ],

                "dara-vrd": [

                "dara-vrd",
```

```
"bortezomib-daratumumab-dexamethasone-lenalidomide"
],
"rd": [
"dexamethasone-lenalidomide",
"rd"
],
"vd": [
"bortezomib-dexamethasone",
"vd"
],
"dvd": [
"bortezomib-daratumumab-dexamethasone",
"dvd"
],
"dara-cybord": [
"dara-cybord",
"bortezomib-cyclophosphamide-daratumumab-dexamethasone"
],
"vzd": [
"dexamethasone-lenalidomide-zometa"
],
"dara-rd": [
"drd rd",
"daratumumab-dexamethasone-lenalidomide"
],
```

```
                "vtd": [

                "vtd",

                "bortezomib-dexamethasone-thalidomide"

                ]

                }}

                NOTES:

                For both the dates, if day is missing then use 1 in place of
dd, if the month and/or year is missing then pick missing value(s) from the
date when note was written which is {note_date} If nothing is clearly
mentioned then output NO_MENTION. DO NOT OUTPUT ANY STRING OTHER THAN
NO_MENTION OR THE DATE IN SPECIFIED FORMAT

                Treatment response labels(Pick most suitable one based on the
IMWG criteria): PARTIAL_RESPONSE, MINIMAL_RESPONSE,
STRINGENT_COMPLETE_RESPONSE, COMPLETE_RESPONSE, VERY_GOOD_PARTIAL_RESPONSE,
NO_RESPONSE, PROGRESSIVE_DISEASE, NO_MENTION # use no mention if there's no
mention of this in the note

                Note that this patient note was written on {note_date} in
case you need to refer to the date of the note.

                ONLY OUTPUT THE JSON AND NOTHING ELSE NO REASONING NEEDED



transplant_status:

        reason: |

                Based on the given patient note:

                {chunk_text}

                Comment on the status of any bone marrow transplant in
context of multiple myeloma, whether the transplant has been done or the
eligibility assessment result is given or if nothing is given in the note,
mention that as well.

                NOTE: Only consider the bone marrow transplant, if the
elgibility assesment etc are in context of some other transplant then ignore

                "Use what's given in the note and do not make any assumptions
or implications."
```

```
        answer: |

                Based on this provide a value for the status of only bone
marrow transplants that may relate to Multiple Myeloma:

                PERFORMED: If the transplant has been done

                ELIGIBLE: If the assessment for bone marrow transplant is
done and doctor has deemed patient eligible for the same

                ELIGIBLE_BUT_DEFERRED: If the patient is eligible but
transplant has been deferred

                NOT_ELIGIBLE: If the patient has been declared ineligible and
advised against taking the transplant

                IN_EVALUATION: "If the above labels aren't applicable and
doctor has only discussed the transplant with patient or if the transplant
eligibility evaluation is not completed"

                NOT_ASSESSED: "If the patient has never been assessed for
bone marrow transplant before or if there's no mention of it in the note"

                Output a json in following format:

                {{"bm_transplant_status": "one of the above labels"}}

                ONLY OUTPUT THE JSON AND NOTHING ELSE NO REASONING NEEDED

        combined: |

                Based on the given patient note:

                {chunk_text}

                Output a value for the status of only bone marrow transplants
that may relate to Multiple Myeloma:

                PERFORMED: If the transplant has been done

                ELIGIBLE: If the assessment for bone marrow transplant is
done and doctor has deemed patient eligible for the same

                ELIGIBLE_BUT_DEFERRED: If the patient is eligible but
transplant has been deferred

                NOT_ELIGIBLE: If the patient has been declared ineligible and
advised against taking the transplant

                IN_EVALUATION: "If the above labels aren't applicable and
```

```
doctor has only discussed the transplant with patient or if the transplant
eligibility evaluation is not completed"

                NOT_ASSESSED: "If the patient has never been assessed for
bone marrow transplant before or if there's no mention of it in the note"

                Output a json in following format:

                {{"bm_transplant_status": "one of the above labels"}}

                ONLY OUTPUT THE JSON AND NOTHING ELSE NO REASONING NEEDED
```
