## Supplementary Note 4 for "Automated abstraction of clinical parameters of multiple myeloma from real-world clinical notes using large language models"

### Supplementary Note 4 - BERT Pipeline Detailed Methodology

The BERT pipeline used as the foundation of the BERT workflow is an assortment of multiple deep neural network models trained to extract relevant information from clinical notes. The models were initially trained on a corpus of 75,000 double-human-annotated sentences extracted from de-identified free text data of the AMC network available in the nference nSights platform. The maximum sequence length for these models is 128 tokens to reduce computational overhead and enable efficient inference.

The information extracted by the BERT pipeline can be divided into two aspects:

1. Entities: All the medically relevant nouns and adjectives present in notes. It is divided into three categories:
    a. Primary Entities: Entities with a proper definition (mostly nouns) like PROBLEM, MEDICINE, DIAGNOSTIC_PROCEDURE…
    b. Secondary Entities: Entities that advise on or provide value to a primary entity. Like VALUE, MED_DOSE, SEVERITY…
    c. Associations: A combination of two entities that are associated with each other.
2. Sentiments: Sentiments are the context a sentence indicates towards the entity/entities present. It can be very subjective. We focus on three different event-based aspects.
    a. Temporal Assessment: Describes the timeline associated with the entity of interest with respect to the sentence
    b. Certainty Assessment: Describes if the entity or an event certainly transpired, given the context of the sentence.
    c. Subject Assessment: Describes whether the entity in the sentence is associated with the patient.

The BERT pipeline consists of the following three model types:

1. Clinical Named Entity Recognition (NER)
2. Sentiment Models
    a. Certainty Assessment Model
    b. Temporality Assessment Model
    c. Subject Classification Model
3. Association Models
    a. Concept Association Model
    b. Generic Date Association Model

#### 4.1 BERT Pipeline Rules

**Supplementary Table 4.1.** BERT pipeline rules by data field

| Variable | Methodology |
|---|---|
| **Multiple Myeloma (MM) diagnosis** | 1. Multiple myeloma mentions were extracted by running the BERT + regex model using the relevant synonyms.<br>2. The extracted sentences were then filtered using the following qualifiers:<br>- Certainty: Yes/Maybe_Yes<br>- Subject: Patient<br>3. The notes were further filtered to remove positive mentions of smoldering myeloma.<br>4. Certainty of 'Yes' was considered as confirmed multiple myeloma, while certainty of 'Maybe_Yes' was considered as a likely case of multiple myeloma. |
| **MM Diagnosis date** | 1. For the confirmed myeloma cases from above, the dates were extracted by one of the following methods:<br>- Extracted date in the previous or subsequent sentence of a confirmed myeloma mention if the term 'diagnosis' (to capture phrases like 'initial diagnosis', 'diagnosis date') is part of the sentence.<br>- For cases not picked up by the above method, the associated dates in the same sentence were considered.<br>2. The earliest date in each note was considered as the date of diagnosis of multiple myeloma. |
| **MM status** | 1. The confirmed myeloma cases were classified into one of the following types:<br>- Newly diagnosed: regex search in the corresponding sentences for relevant phrases (e.g.: "recent diagnosis of multiple myeloma") with temporality - 'Current' (or) the diagnosis date (extracted from above) within 3 months prior to the note date<br>- Relapse: regex search in the corresponding sentences for relevant phrases (e.g.: "relapsed multiple myeloma") with temporality - 'Current'<br>- Remission: regex search in the corresponding sentences for relevant phrases (e.g.: "multiple myeloma in remission") with temporality - 'Current' |
| **MM-related bone lesion presence** | 1. Bone lesions were extracted by running the BERT + regex model using the relevant synonyms.<br>2. The extracted sentences were then filtered using the following qualifiers:<br>- Certainty: Yes |

| | |
|---|---|
| | - Subject: Patient<br>- Temporality:<br>- Current<br>- Any temporality If there is an associated date which is within 6 months prior to the note date<br>3. The sentences were further filtered to remove mention of lesions not involving the bone. |
| **ECOG Score** | 1. ECOG mentions were extracted by running the BERT + regex model using the relevant synonyms.<br>2. The extracted sentences were then filtered using the following qualifiers:<br>- Certainty: Yes<br>- Subject: Patient<br>- Temporality:<br>- Current<br>- Any temporality If there is an associated date<br>3. The ECOG values were filtered to ensure they were within limits (0-4), and the most recent value in each note was considered. |
| **EMD presence** | 1. Extramedullary disease was extracted by running the BERT + regex model using the relevant synonyms.<br>2. The extracted sentences were then filtered using the following qualifiers:<br>- Certainty: Yes<br>- Subject: Patient<br>- Temporality:<br>- Current<br>- Any temporality If there is an associated date which is within 6 months prior to the note date |
| **EMD location** | 1. The associated anatomical structures from the above extramedullary disease mentions were harmonized into soft tissue and paramedullary sites, and then compared with the ground truth. |
| **Plasmacytosis percentage** | 1. Plasmacytosis percentage was extracted by running the BERT + regex model using the relevant synonyms.<br>2. The extracted sentences were then filtered using the following qualifiers:<br>- Certainty: Yes<br>- Subject: Patient<br>- Temporality:<br>- Current<br>- Any temporality If there is an associated date |

| | |
|---|---|
| | 3. The plasmacytosis values were filtered to ensure they were within limits (0-100), values with irrelevant units (e.g.: ‘mg’, ’g/L’, ’cells/million’) were removed, and the most recent value in each note was considered. |
| **Transplant eligibility & status** | 1. Transplant mentions were extracted by running the BERT + regex model using the relevant synonyms.<br>2. The extracted sentences were then filtered using the following qualifiers for each transplant status:<br>- Performed:<br> - Certainty: Yes<br> - Subject: Patient<br> - Temporality:<br> - History<br> - Current_Uncertain if transplant date on or before note date<br>- Eligible:<br> - Certainty: Yes<br> - Subject: Patient<br> - Temporality: Upcoming<br>- Not eligible:<br> - Certainty: No<br> - Subject: Patient |
| **FLT response** | 1. The treatment responses (CR, sCR, VGPR, PR, PD) were extracted using the BERT + regex model using the relevant synonyms.<br>2. The extracted sentences were then filtered using the following qualifiers:<br>- Yes, Patient with any temporal sentiment if there is an associated date<br>- Yes, Patient, Current/Current_Uncertain/History if there is no associated date (note date kept as associated date)<br>3. The sentences with irrelevant associated anatomical structures (for e.g.: lower back, MCA) are removed.<br>4. The earliest response for each note is retained and considered as the response to the first line of therapy. |
| **FLT response date** | 1. From the above final set of first line therapy response extractions, those with an extracted date associated with the response or those with a ‘Current’ temporality for the extracted response are alone filtered and considered as the date of first line therapy response. |
| **FLT regimen** | 1. Two synthesizer instances to capture LOT:<br>- Line of therapy (Procedure) with relevant synonyms of drug regimens<br>- All medications |

| | |
|---|---|
| | 2. The following extractions were filtered out from both LOT instance and all medications instance:<br>- Yes, Patient with any temporal sentiment if there is an associated date<br>- Yes, Patient, Current/Current_uncertain/History if there is no associated date (note date kept as associated date)<br>3. For notes where the LOT instance is not available after the above filters, the relevant medications (MM drugs) are filtered out from all medications instance and grouped by sentence, variable date and temporality to form the regimen.<br>4. Both the final filtered extractions from the LOT instance and all medications instances are combined and the earliest regimen for each note is retained and considered as the first line of therapy. |
| **FLT start date** | 1. From the above final set of first line therapy extractions, those with an extracted date associated with the drugs/regimen are alone filtered and considered as the date of start of first line therapy. |

#### 4.2 BERT+Regex Model Metrics:

**Supplementary Table 4.2a.** Named entity recognition (NER) model performance

NER (Named entity Recognition) is a Deep Neural Network model that given a clinical sentence, can extract all the relevant biomedical entities with their relevant tags.

| S. No | Label | Precision | Recall | F1 |
|---|---|---|---|---|
| 1. | PROBLEM | 0.85 | 0.86 | 0.85 |
| 2. | LAB_DATA | 0.90 | 0.91 | 0.91 |
| 3. | MEDICAL_SURGICAL_PROCEDURE | 0.79 | 0.80 | 0.79 |
| 4. | LAB_PROCEDURE | 0.80 | 0.85 | 0.82 |
| 5. | DIAGNOSTIC_PROCEDURE | 0.85 | 0.84 | 0.84 |
| 6. | MEDICINE | 0.90 | 0.92 | 0.91 |
| 7. | ANATOMICAL_STRUCTURE | 0.80 | 0.82 | 0.81 |
| 8. | BODY_FUNCTION | 0.57 | 0.62 | 0.59 |
| 9. | BODY_MEASUREMENT | 0.85 | 0.87 | 0.86 |
| 10. | CHRONOLOGY | 0.90 | 0.92 | 0.91 |
| 11. | MEDICAL_DEVICE | 0.79 | 0.85 | 0.82 |
| 12. | MED_DOSE | 0.85 | 0.87 | 0.86 |
| 13. | MED_DURATION | 0.77 | 0.72 | 0.75 |
| 14. | MED_FORM | 0.89 | 0.89 | 0.89 |
| 15. | MED_FREQUENCY | 0.87 | 0.87 | 0.87 |
| 16. | MED_ROUTE | 0.88 | 0.89 | 0.88 |
| 17. | MED_STATUS | 0.76 | 0.82 | 0.79 |

| | | | | |
|---|---|---|---|---|
| 18. | MED_STRENGTH | 0.83 | 0.7 | 0.85 |
| 19. | MED_TOTALDOSE | 0.68 | 0.73 | 0.70 |
| 20. | MED_UNIT | 0.96 | 0.90 | 0.93 |
| 21. | NORMAL_FINDING | 0.71 | 0.72 | 0.72 |
| 22. | PATIENT_STATUS | 0.53 | 0.64 | 0.58 |
| 23. | RESULT | 0.75 | 0.81 | 0.78 |
| 24. | SEVERITY | 0.8 | 0.81 | 0.8 |
| 25. | SUBSTANCE_ABUSE | 0.73 | 0.86 | 0.79 |
| 26. | UNIT | 0.90 | 0.90 | 0.90 |
| 27. | VALUE | 0.92 | 0.93 | 0.92 |

**Supplementary Table 4.2b.** Certainty assessment model performance

Given a sentence and a target entity present in that sentence, the certainty model predicts the certainty aspect of the entity.

| Labels | Precision | Recall | F1 |
|---|---|---|---|
| CONDITIONAL | 0.67 | 0.49 | 0.56 |
| MAYBE_NO | 0.47 | 0.44 | 0.45 |
| MAYBE_YES | 0.71 | 0.64 | 0.68 |
| NO | 0.88 | 0.89 | 0.89 |
| NO_CERTAINTY | 0.64 | 0.51 | 0.57 |
| UNCERTAIN | 0.51 | 0.41 | 0.45 |
| YES | 0.94 | 0.97 | 0.96 |

**Supplementary Table 4.2c.** Temporal assessment model performance

Determining whether an event occurred in the past, present, or has the potential to happen in the future can provide valuable insights into the sentence's context. This is handled by the temporal assessment model.

| Labels | Precision | Recall | F1 |
|---|---|---|---|
| CURRENT | 0.86 | 0.87 | 0.86 |
| CURRENT_UNCERTAIN | 0.56 | 0.51 | 0.53 |
| HISTORY | 0.74 | 0.8 | 0.77 |
| NO_TEMPORAL_ASPECT | 0.64 | 0.52 | 0.57 |

| | | | |
|---|---|---|---|
| UPCOMING | 0.75 | 0.73 | 0.74 |

**Supplementary Table 4.2d.** Subject assessment model performance

The model we use is designed to extract whether the entity or event mentioned in a sentence is associated with the patient or not. This can be crucial in determining the relevance of the information to the patient's current medical situation.

| Labels | Precision | Recall | F1 |
|---|---|---|---|
| BLOOD_RELATIVE | 0.87 | 0.91 | 0.89 |
| NO_SUBJECT | 0.65 | 0.52 | 0.58 |
| PATIENT | 0.97 | 0.98 | 0.98 |

**Supplementary Table 4.2e.** Association model performance

In a sentence, simply extracting entities may not be sufficient since many of them only convey meaning when they are linked to another entity present in the sentence. Therefore, it is crucial to extract the relationships between all entities to fully capture the meaning conveyed by the sentence. This is handled by the association model.

| Pair Type | F1_Yes | F1_No |
|---|---|---|
| LAB_DATA___VALUE | 0.99 | 0.999 |
| UNIT___VALUE | 0.99 | 0.998 |
| ANATOMICAL_STRUCTURE___PROBLEM | 0.97 | 0.98 |
| BODY_MEASUREMENT___VALUE | 0.997 | 0 |
| ANATOMICAL_STRUCTURE___MEDICAL_SURGICAL_PROCEDURE | 0.92 | 0.98 |
