## Supplementary Note 5 for "Automated abstraction of clinical parameters of multiple myeloma from real-world clinical notes using large language models"

### Supplementary Note 5 - LLM Sensitivity Analysis

Temperature and top-k hyperparameter settings were varied to explore the robustness of the prompts and prompting techniques used for extraction.

For hyperparameter sensitivity analyses, the best-performing Llama workflows were run with 6 pairs of temperature (values of 10, 50) and top-k (values of 0.4, 0.7 and 1.0) values. For each pair, the experiment was repeated 10 times with different seeds. The resulting F1-scores from these 10 runs were averaged to estimate the variance in accuracy and to assess the overall robustness of the LLM workflows used for each clinical concept.

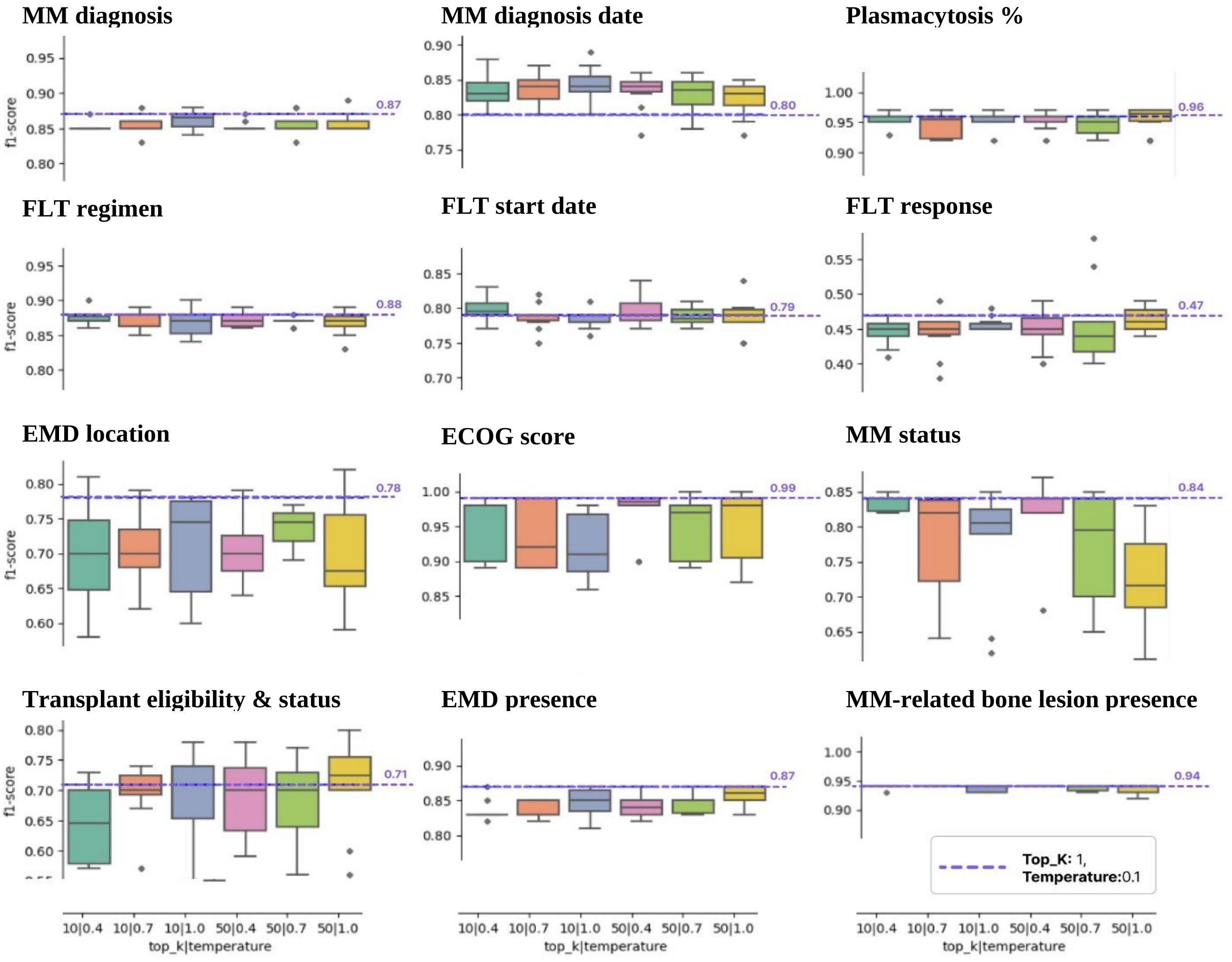


**Supplementary Figure 5:** Performance variance over 10 iterations with random seeds for different combinations of temperature and top k values for the best-performing LLM workflow in the test set. The purple dotted line indicates the performance with top-k=1 and temperature=0.1
